# Evidence from Madagascar shows that vaccination could mitigate climate-driven disruptions to malaria control

**DOI:** 10.1101/2024.04.02.24305230

**Authors:** Benjamin L. Rice, Estelle Raobson, Sylviane Miharisoa, Mahery Rebaliha, Joseph Lewinski, Hanitriniaina Raharinirina, Christopher D. Golden, Gabriel A. Vecchi, Amy Wesolowski, Bryan Grenfell, C. Jessica E. Metcalf

## Abstract

Extreme weather events are common in high malaria burden areas and likely to increase in severity due to climate change. Yet, data on infection rates following these events and the consequences for disease control programs remain rare. Using data from Madagascar, we estimate high rates of infection in the wake of two major tropical cyclones and show infection rebounds rapidly during gaps in standard interventions. Relative to other control options, recently available malaria vaccines have a longer duration of protection, with the potential to address interruptions in prevention deployment. Evaluating this use, we quantify the reduction in symptomatic infections expected for a range of vaccination scenarios. We find long-lasting interventions such as vaccination are a key mitigation measure against climatic disruptions to disease control.

## Main Text

Malaria remains a major global health challenge (*1*, *2*). Much work has investigated how the coincident challenge of climate change threatens progress to malaria control (*1*). Rising temperatures modify vector mosquito dynamics (*3–5*), altering infection risk (*6*, *7*), but these impacts are often focused in low burden areas at the altitudinal and latitudinal margins of malaria transmission (*8–11*). By contrast, extreme weather events, some types of which are likely to increase in frequency or severity due to climate change (*12–15*), disrupt public health efforts (*16–21*), including in areas where the burden of malaria is concentrated. Data on the impact of climate-related disasters on malaria infection remain rare (*19*, *21–24*), despite high rates of exposure to such events in malaria endemic settings.

Evaluating the malaria burden associated with such disruptions and potential mitigation strategies requires considering the changing landscape of malaria control activities. In response to stalled progress in malaria control in high burden countries and the development of new treatment and prevention tools (*1*, *25*), the World Health Organization (WHO) and national malaria control programs are preparing to scale the deployment of new interventions, including the mass distribution of antimalarial chemoprophylaxis and treatment (e.g., seasonal or perennial malaria chemoprophylaxis, SMC, PMC), and vaccination (see **Table S1**). Critically, continuity is important to these interventions, as their effectiveness erodes over time, thus requiring regular re-administration. Extreme weather events threaten that continuity by delaying or degrading public health activities (*19*). Understanding the barrier this poses to achieving malaria control targets requires detailed data on infection risk during periods of likely disruption. However, limitations in surveillance systems make such data rare.

Tropical cyclones are an example of extreme weather events that disrupt health systems for weeks to months following the event (*23*, *26–30*). Madagascar, our focus here, is among numerous malaria endemic countries subject to regular cyclones (**Figure 1**), resulting in widespread destruction, population displacement, and damage to healthcare-related infrastructure (**Data S1**). Madagascar is also vulnerable to climate change including from the increasing frequency and severity of major cyclones in the southern Indian Ocean (*31*, *32*) with storms causing repeated humanitarian emergencies in recent years. Cyclones Batsirai and Freddy destroyed over 300 health facilities (*33–35*) and resulted in an estimated 112,115 and 290,000 people in need of immediate humanitarian assistance in 2022 and 2023, respectively (*36*) (**Figure 1D**). Excess rainfall for Cyclone Batsirai was made more likely by increased greenhouse gas and changes in aerosol emission per climate attribution studies (*37*).

**Figure 1:**
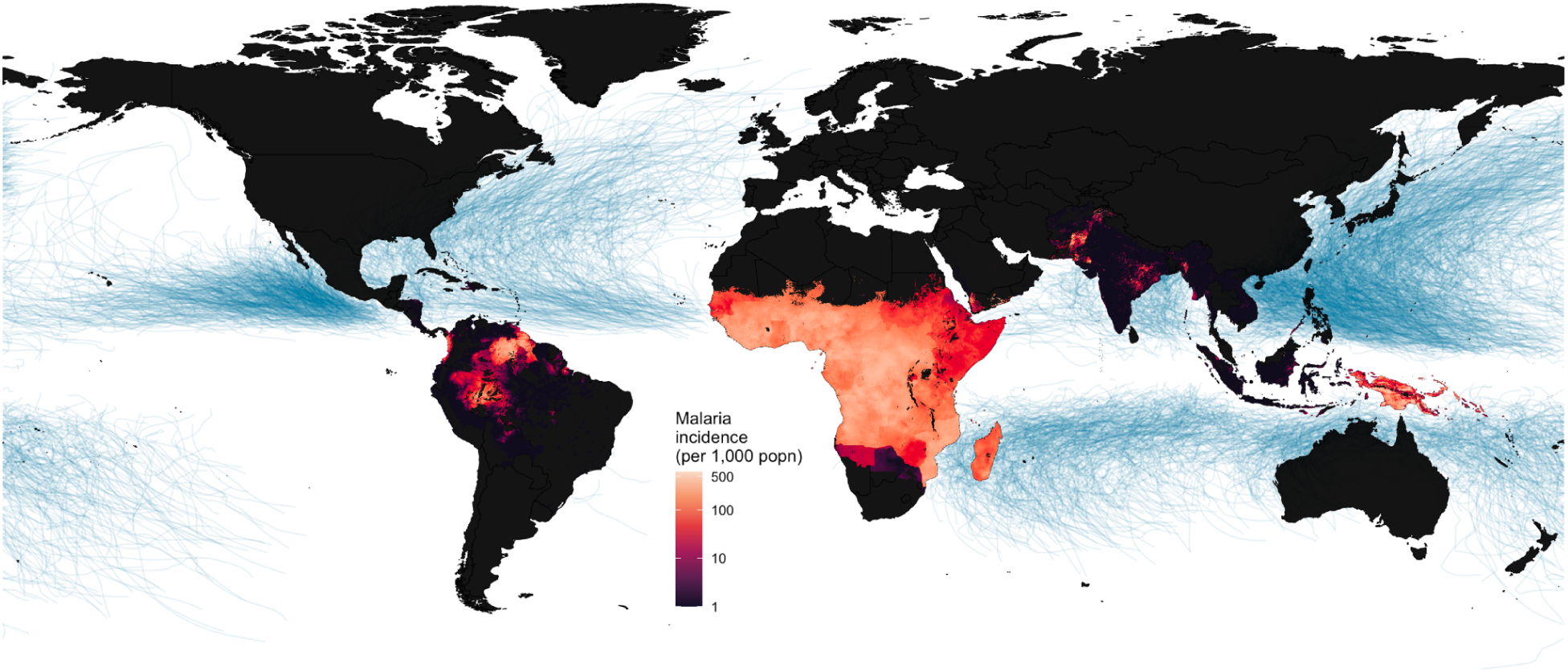
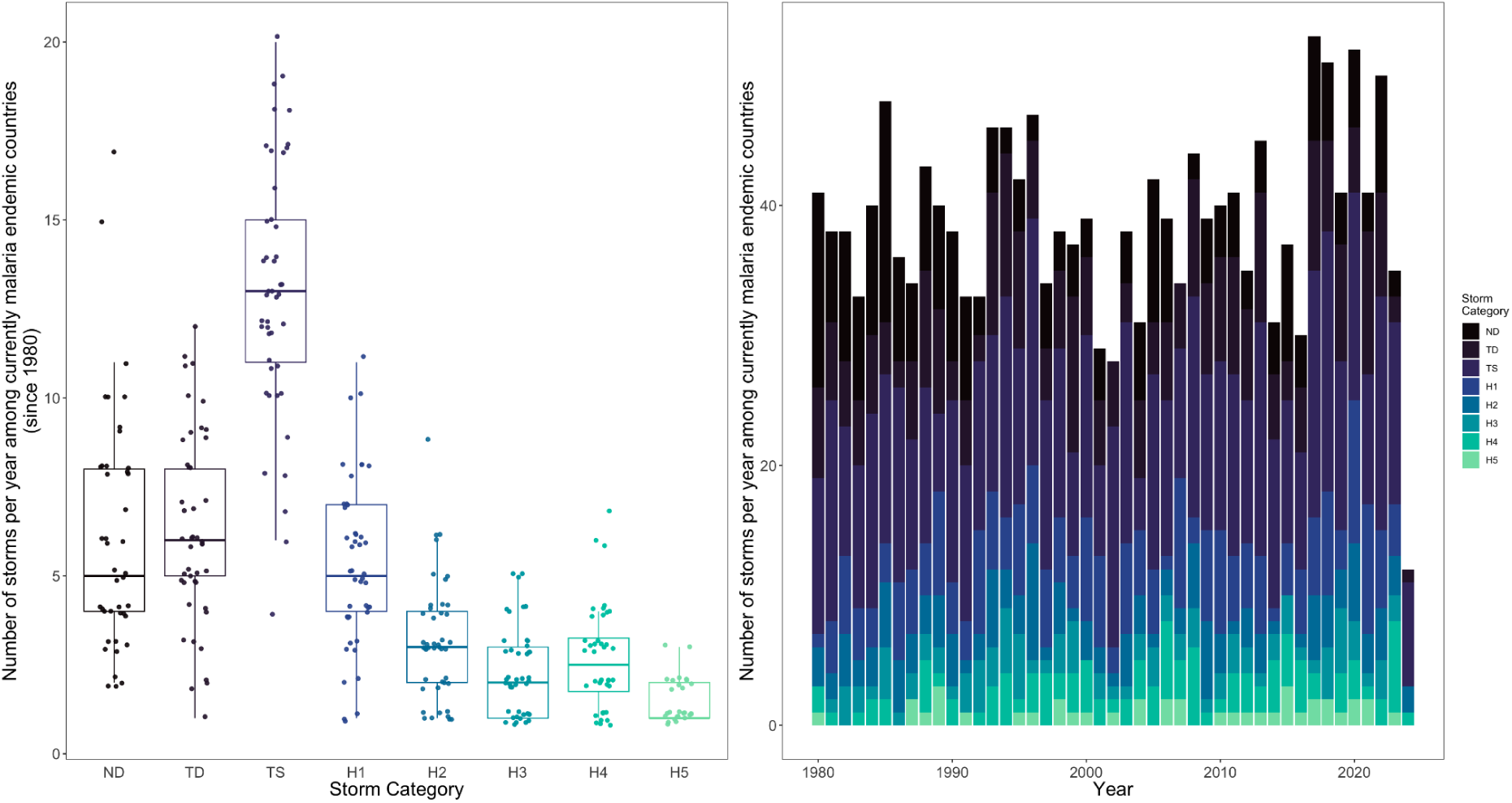
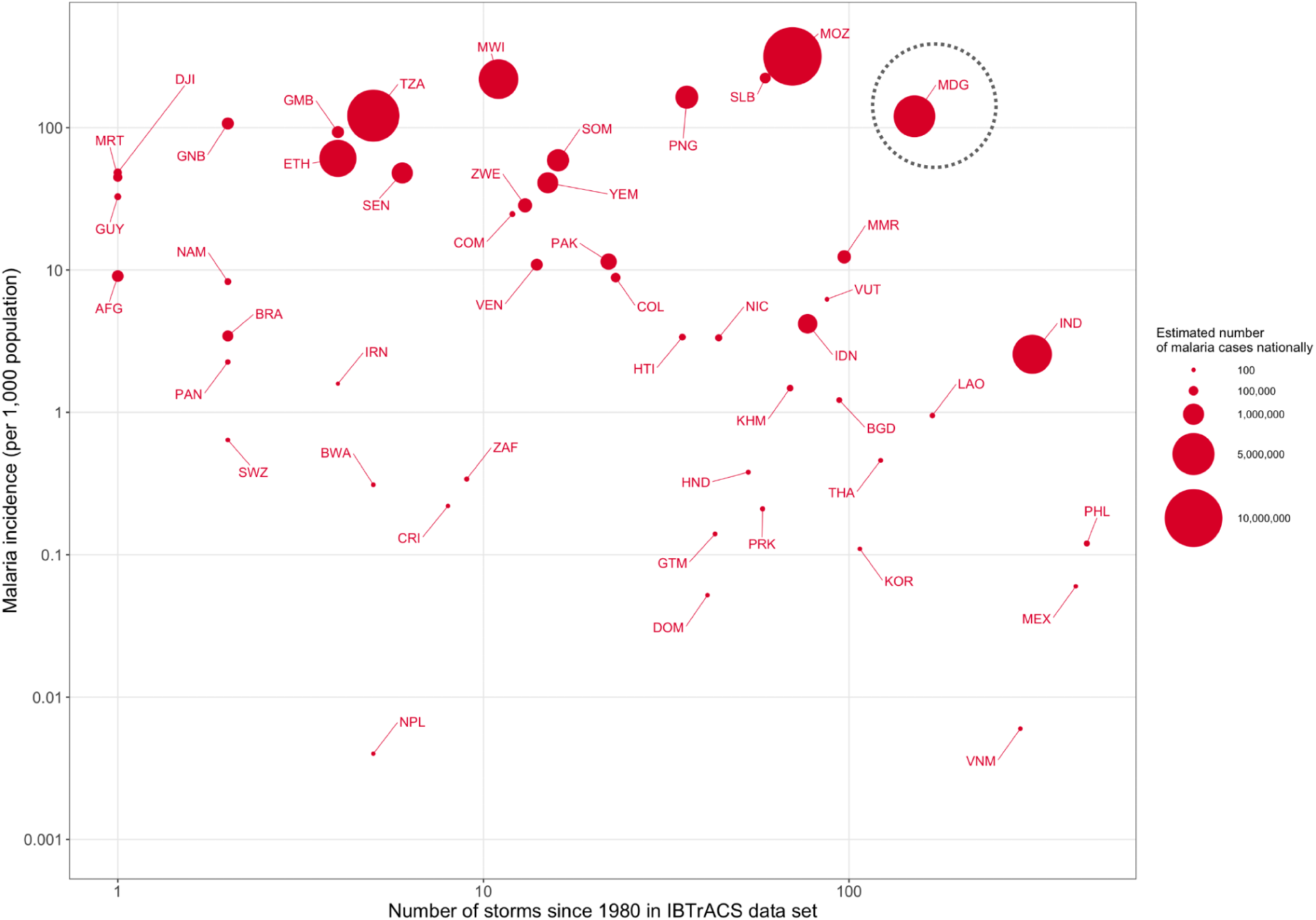
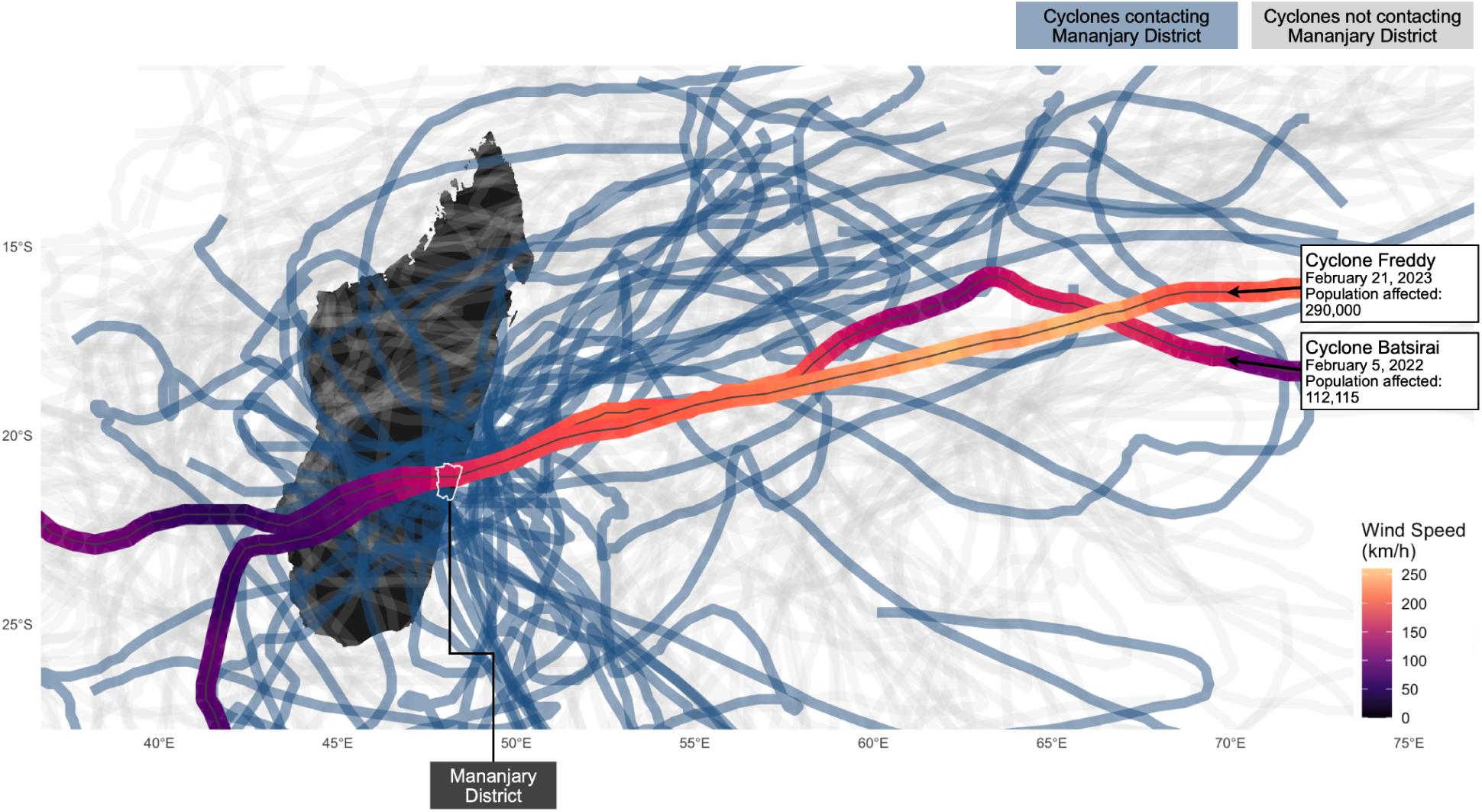
The scale of cyclone-malaria interactions globally and in Madagascar. **Figure 1A**: Blue lines show cyclone tracks from the International Best Track Archive for Climate Stewardship (IBTrACS) for all storms since 1980 (*38*, *39*). Malaria incidence estimates are from the Malaria Atlas Project data for 2022 (*40*). **Figure 1B**: Intensity for the 1897 recorded storms contacting currently malaria endemic countries since 1980 (defined as storm center approaching within 60 nautical miles of landfall). For boxplots, the midline shows the median number of storms in the period 1980-2024, lower and upper hinges correspond to the 25th and 75th percentiles and whiskers extend 1.5 times the interquartile range. Points show the number of storms for a given year, jittered for visibility. Categories follow the Saffir-Simpson wind scale for the maximum wind speed observed within 60 nautical miles of landfall: Tropical Depression (TD), Tropical Storm (TS), Hurricane Category 1-5 equivalent (H1-H5), or no wind speed data available (ND). **Figure 1C**: Malaria incidence vs the number of tropical cyclones since 1980 passing within a 60 nautical mile buffer of a country’s border (from IBTrACS), on a log-10 scale. Point size shows the estimated number of malaria cases contributed per country (most recent data from 2021). Malaria incidence and case data are from the World Health Organization (WHO) Global Health Observatory. Points are labeled by three-letter country codes (countries with no reported malaria cases or no observed cyclones not shown). Among higher malaria burden countries (annual incidence > 100 per 1000 population), the highest cyclone frequencies were observed for Madagascar (MDG, circled), Mozambique (MOZ), the Solomon Islands (SLB), Papua New Guinea (PNG), Malawi (MWI) and Tanzania (TZA). **Figure 1D**: Tropical cyclone activity for Madagascar since 1980 with the study district, Mananjary, outlined. Tracks show the estimated path of the storm center, with those passing within 60 nautical miles (111.1 km) of the Mananjary district shown in blue (others in gray). Two examples of recent cyclones impacting the Mananjary district, Cyclone Batsirai and Cyclone Freddy, are highlighted with estimated wind speed (data from IBTrACS) and estimates of the number of individuals requiring humanitarian assistance (data from EM-DAT).

Here, we leverage a dataset comprising repeated measures of malaria infection status surrounding cyclone events (*n* = 20,718 observations) from Mananjary district, a moderate to high malaria transmission area located on the east coast of Madagascar, to develop a platform to explore the spectrum of recommended malaria interventions and their robustness under extreme events associated with climate change, providing estimates relevant to a wide range of cyclone prone countries where malaria burden is moderate to high (**Figure 1**, **Figure S1**).

## Results

### 1. High variation in rates of malaria infection following cyclones

#### 1.1 Characterizing malaria infection rates

We use a longitudinal cohort study of a random sample of 500 households (2954 individuals, all ages) from 10 localities (**Figure 2A**). Repeated sampling of malaria infection status by rapid diagnostic test (RDT) was performed for the period July 2021-April 2023 (mean number of samples per enrolled individual = 7.7, median = 9) (**Figure S2**). The interval between samples was short (mean = 57.4 days, median = 52 days) relative to the duration of an untreated *P. falciparum* infection (approx. 180 days (*41*, *42*)), indicating a low probability of natural infection clearance prior to the next sample. All individuals positive for malaria were treated to clear infections, allowing estimates of rates of infection and re-infection (see methods). During the course of the study, the area was impacted by Cyclone Batsirai (February 5, 2022) and Cyclone Freddy (February 21, 2023), providing estimates of malaria infection rates in the period before and for the 2 months after each event (Figure S3).

**Figure 2:**
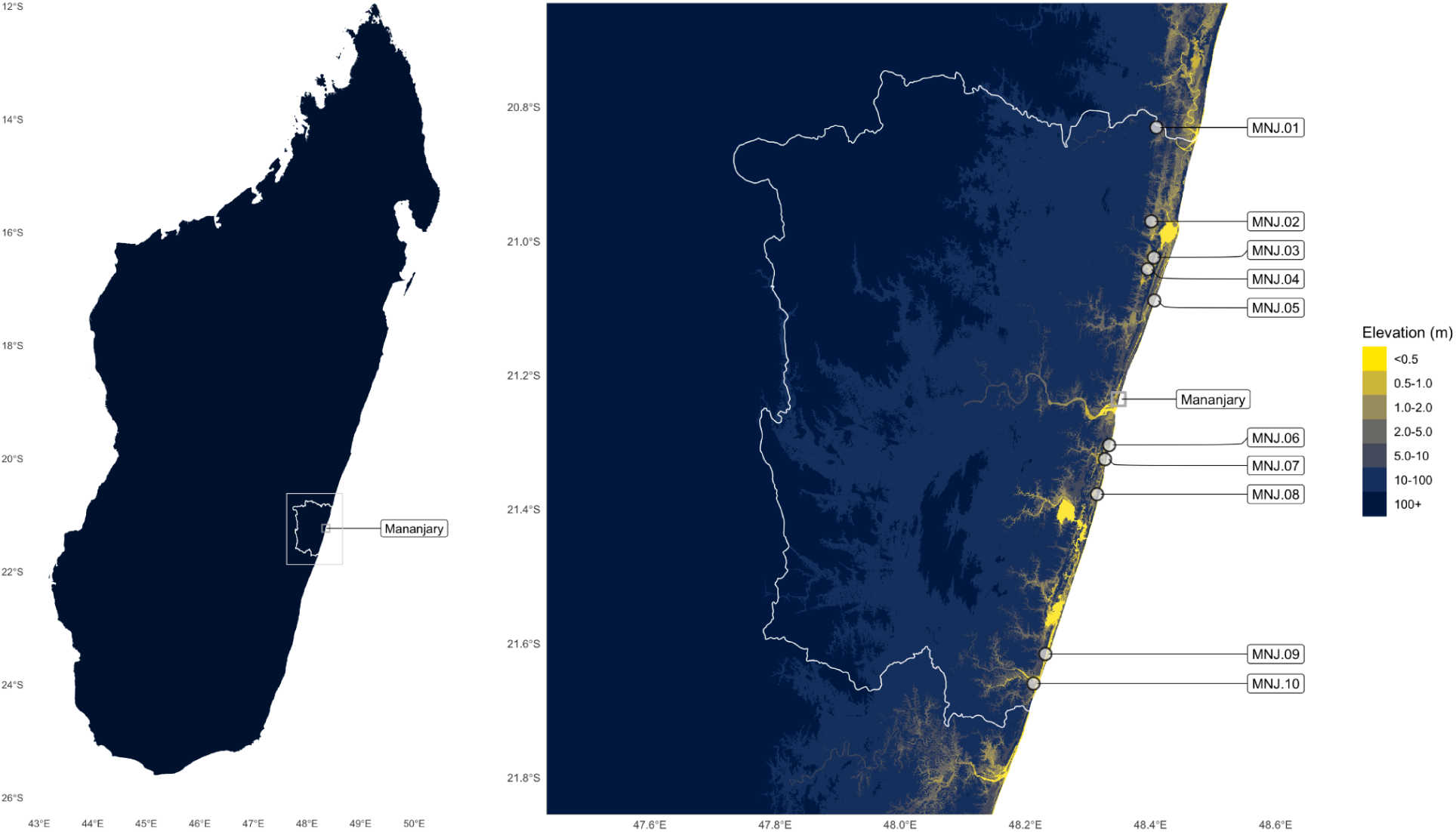
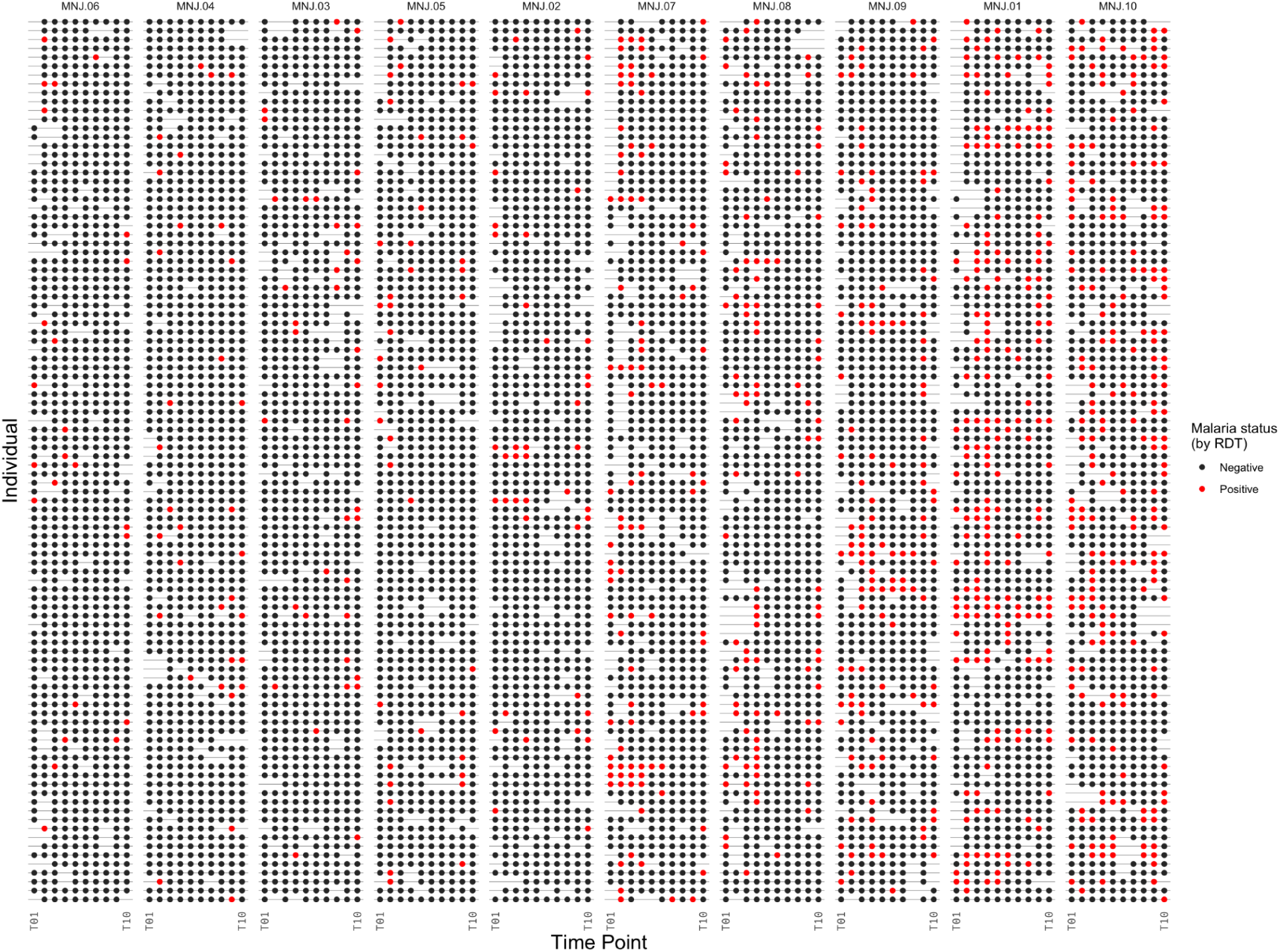
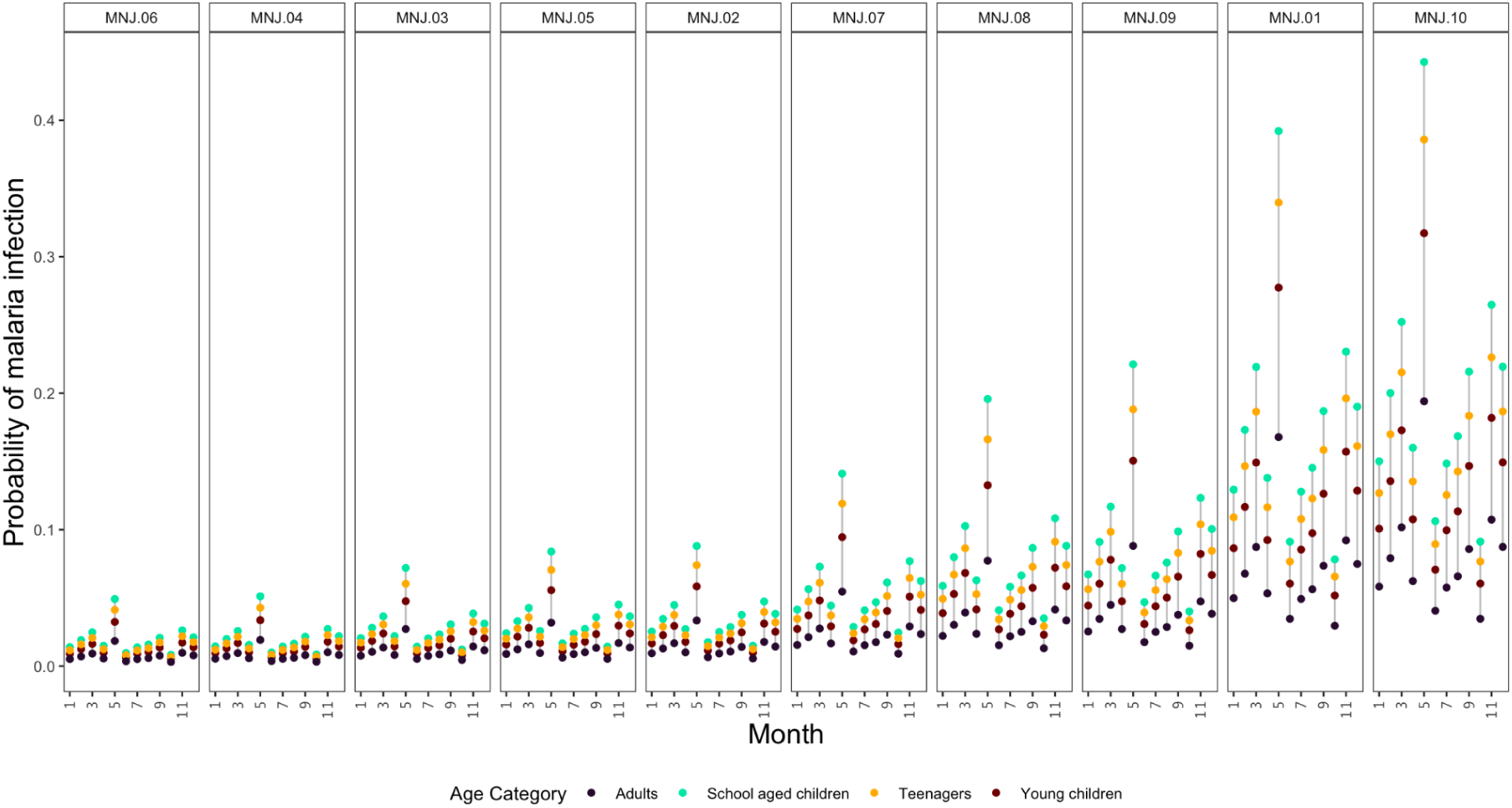
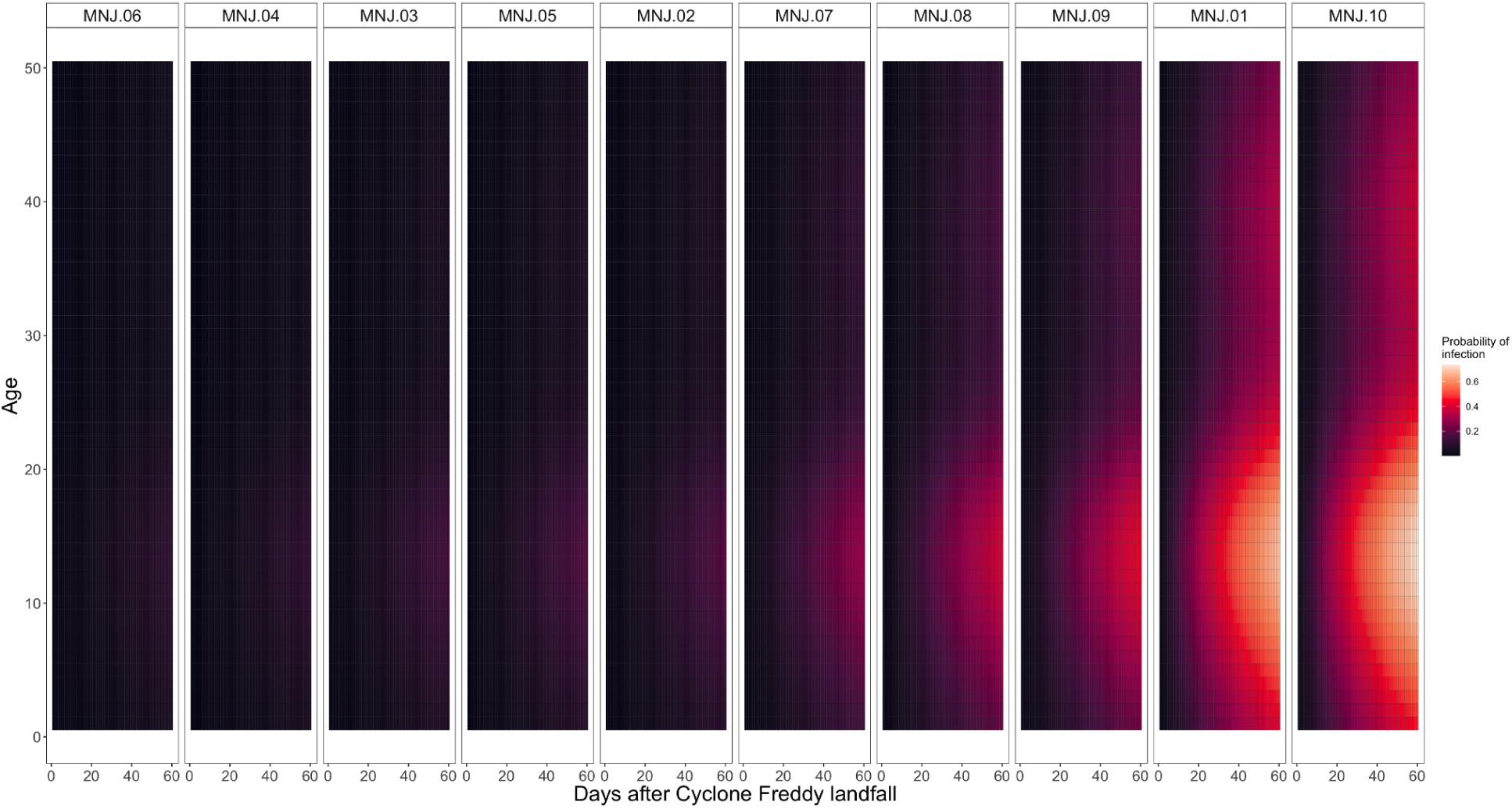
Malaria infection rates following tropical cyclones in southeast Madagascar. **Figure 2A**: Sampling locations for cohort study sites (MNJ.01-MNJ.10, ordered from north to south). Location of the Mananjary urban area, the capital of the Mananjary district, shown with a square. District boundary shown in white, elevation data from the Copernicus Global Digital Elevation Model (*43*). **Figure 2B**: Raw malaria infection observations by rapid diagnostic test (RDT). Individual observations are shown as dots, colored red when positive for malaria. Note RDT positive individuals were treated with antimalarials to clear infections and estimate time to next infection. For visualization, each row is an individual with a random subsample of 100 individuals shown for each locality (individuals with >50% of attendance at sampling time points shown, full data in supplement). The x-axis shows sampling time points for the 10 sampling time points (T01-T10). Panels show the 10 sample localities in Mananjary District, Madagascar (MNJ.01-MNJ.10), ordered from lowest to highest average rate of malaria infection. **Figure 2C**: Expected proportion of individuals infected in one month by site and age category (young children = 0-5y, school-aged children = 6-13y, teenagers = 14-19y, adults = 20+y). Sites ordered by infection rate. **Figure 2D**: Estimated probability of a new malaria infection detectable by rapid diagnostic test (RDT) after Cyclone Freddy landfall (February 21, 2023) at study sites in Mananjary district, Madagascar. Sites ordered by infection rate.

#### 1.2 Force of malaria infection following cyclones

We derive estimates of the force of infection (FOI) by age and season from ten follow-up sampling time points (see methods) (**Figure 2B**). FOI peaked for school-aged children (**Figure S4**), consistent with the higher frequency of infection commonly observed in this group (e.g., (*44*, *45*)). Transmission fluctuated seasonally, with the percentage of school-aged children expected to be infected within a month ranging, by site, from 0.8-9.1% in October to 4.9-44.3% in May (**Figure 2C**). At the higher FOI sites (e.g., MNJ.01, MNJ.08, MNJ.09, MNJ.10), we estimate 13.5-35.6% of younger children and 20.0-49.1% of school aged children were infected with malaria in the 2 months following Cyclones Batsirai and Freddy (**Figure 2D**). The age structure of infection was similar across sites and similar before and after the cyclones (Figure S3). Equipped with these empirically derived estimates of the scale of the rate of infection, we can evaluate the potential impacts of disruptions to malaria control programs during cyclone season.

### 2. Malaria incidence is sensitive to brief disruptions in control

#### 2.1 Consequences of temporal gaps in malaria intervention coverage

Current operational plans for high malaria burden countries, including Madagascar (*46*), recommend implementing or expanding interventions using anti-malaria drugs to reduce burden in targeted populations, as a supplement to existing base activities (i.e., bednet distribution and routine diagnosis and treatment). Approaches use the regular administration of either a standard first-line antimalarial with a short half-life, protective against reinfection for 13-15 days (*47*) to clear existing infections (mass drug administration, MDA, or mass testing and treatment, MTaT (*48*)) or a longer lasting antimalarial, protective for 21-42 days (*49*, *50*) to also prevent new infections (seasonal or perennial malaria chemoprophylaxis, SMC, PMC, or intermittent preventive treatment of malaria, IPT) (see **Table S1**).

We explore the deployment of each of these supplemental interventions across the observed range of FOIs, on the background of existing access to routine clinical care, introducing delays to follow-up administration rounds at different points along a seasonal cycle (see methods). Following the cessation of drug administration, protection decays at a rate determined by the drug half-life, and infections accumulate at a rate determined by the FOI for that locality, age group, and seasonal period (**Figure 3A**). Historical cyclone data for Madagascar indicates the time interval with the highest risk for storm impact occurs between January-April, contributing 81.8% of recorded tropical cyclones (**Figure 3B**). For a disruption that results in prophylactic protection ceasing on March 1st, we estimate 5.3-11.9% of younger children and 8.0-17.6% of school aged children are infected by day 20 in high FOI sites. For comparison, assuming the observed FOI and an efficacy against infection of 88% for prophylaxis (*51*)Given infection prevalence quickly exceeds the WHO defined thresholds for low burden (1-10%), this indicates that the temporary gaps in prophylaxis administration likely in the aftermath of extreme weather events are a major challenge for control programs aiming to maintain burden in the low or near elimination phase.

**Figure 3:**
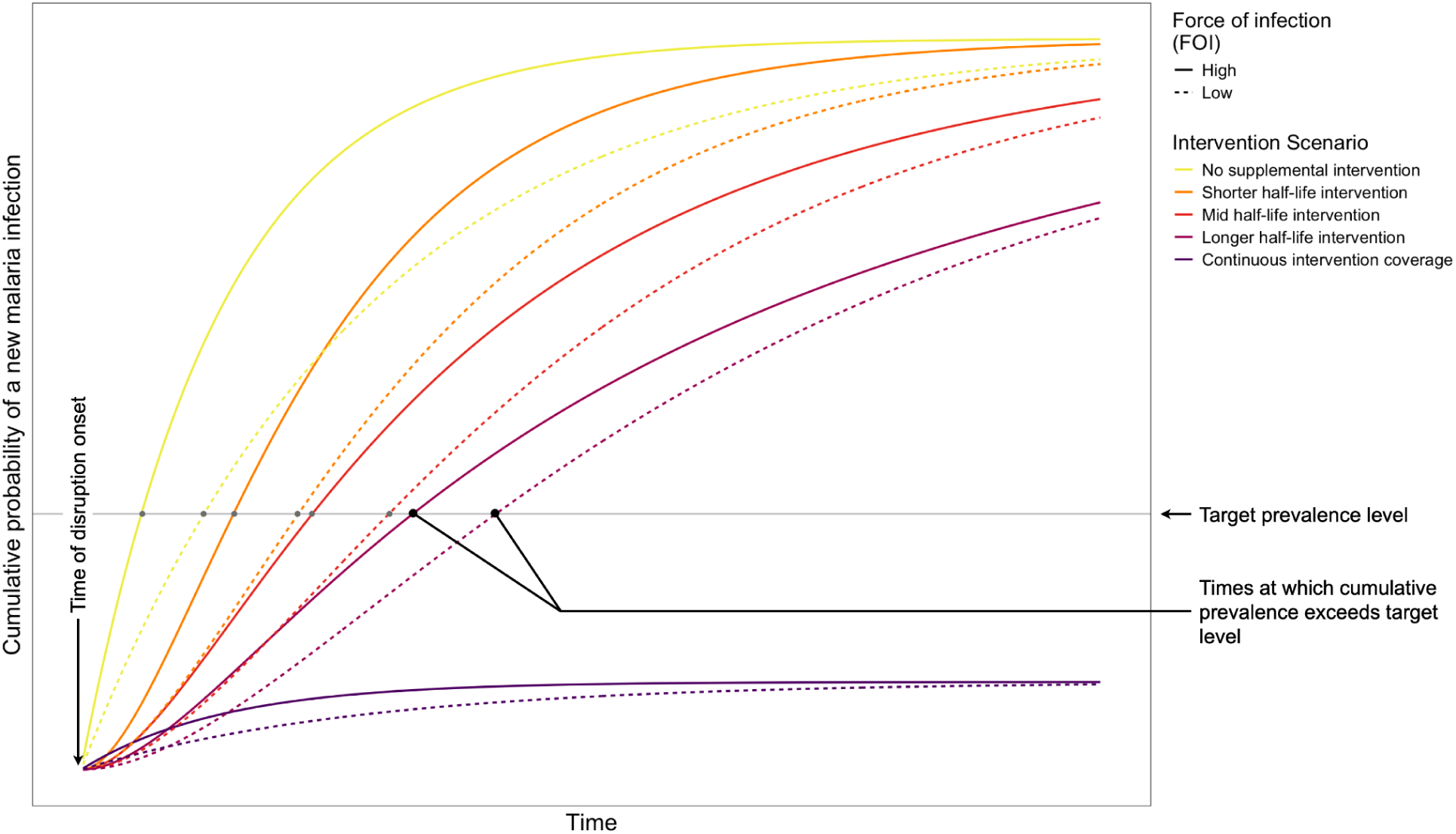
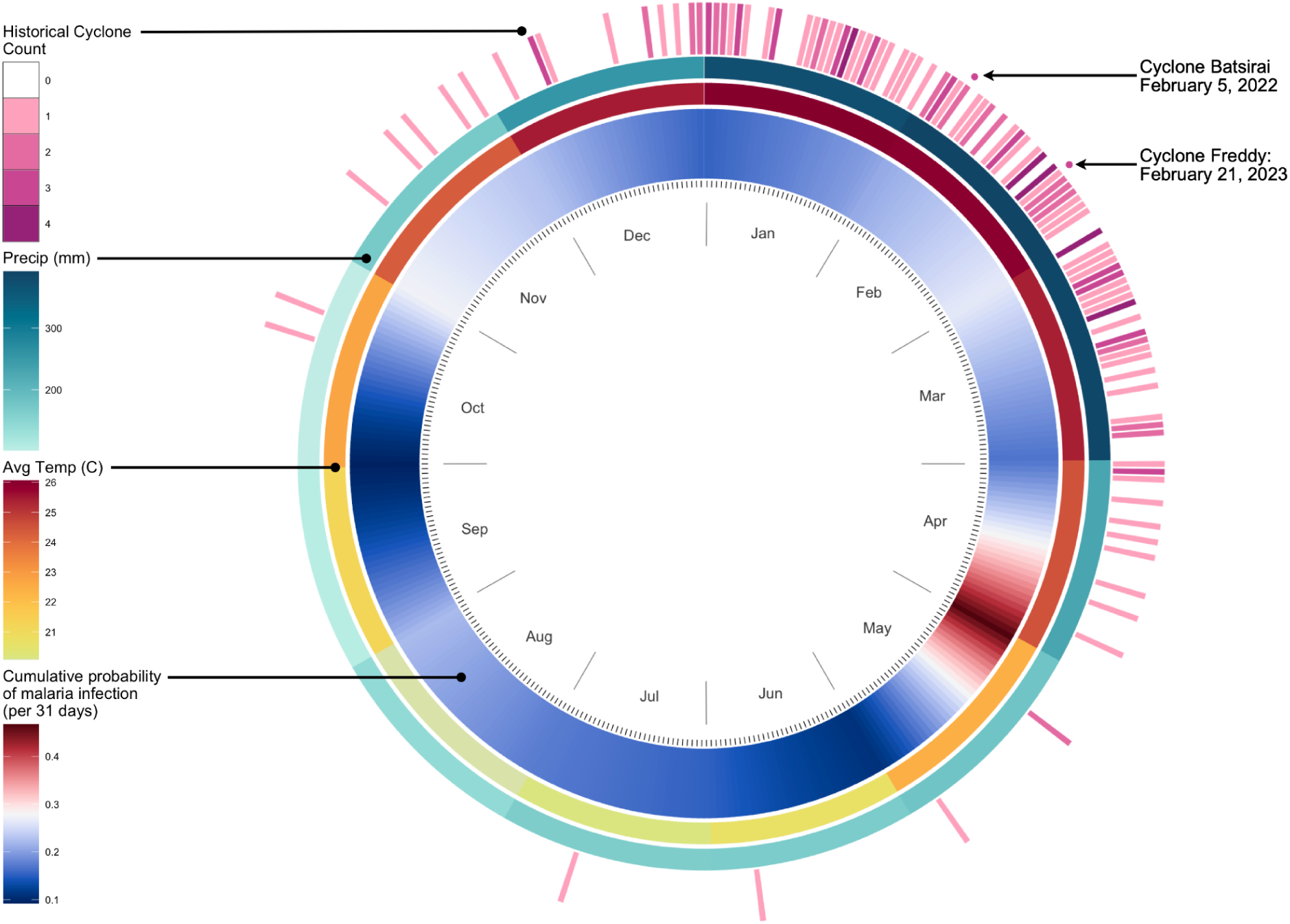
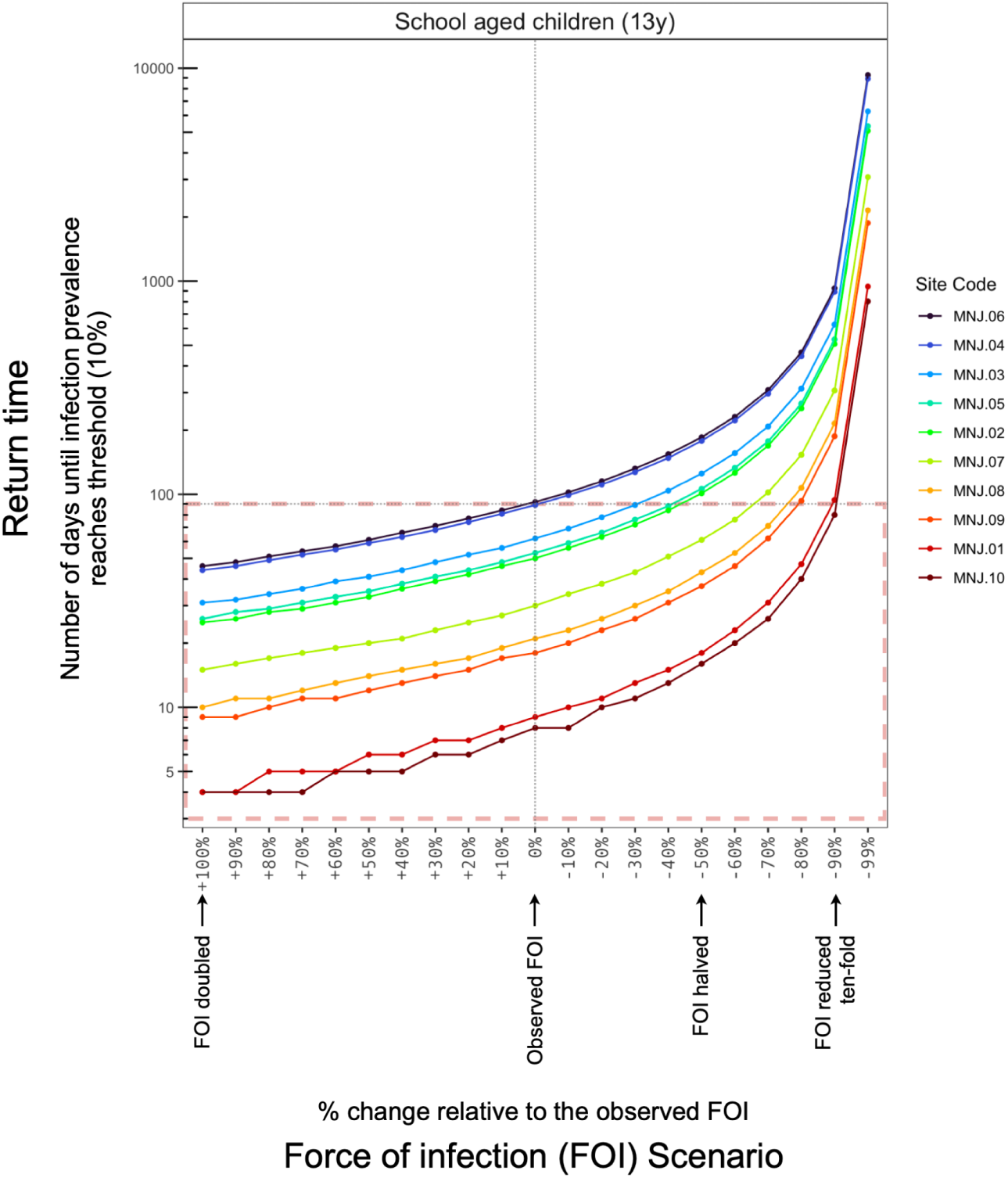
Modeling increases in malaria infection due to interrupted interventions. **Figure 3A**: A schematic reflecting the risk of malaria infection over time as a function of the force of infection (FOI, line types) and the rate of decay of protection from interventions such as chemoprophylaxis. Scenarios show the cumulative probability of acquiring one or more new malaria infections over time: (i) in the absence of any supplemental interventions, (ii-iv) for a population that experiences a halt in the distribution of an intervention with a short, moderate, or long half-life, or (v) continuous, uninterrupted coverage of an intervention (e.g., seasonal/perennial malaria chemoprophylaxis, S/PMC), assuming implementation is in place at the time of disruption for ease of comparison. For illustration, prophylaxis effectiveness estimates are taken from estimates of the protection against infection over monthly intervals for SMC programs (*51*). Results from two FOI estimates are shown, corresponding to the observed FOI for children at localities MNJ.07 (higher FOI) and MNJ.03 (lower FOI). **Figure 3B**: Outer ring: Calendar date of recorded tropical storms impacting Madagascar since 1980. Second and third rings: Average monthly precipitation and temperature for Mananjary district (data from WorldClim 2.1). Inner ring: The cumulative probability of malaria infection (i.e., the proportion of susceptible individuals expected to have one or more new RDT detectable infections) for a 31-day interval beginning on that day. Data shown for school aged children (14 years of age) for a high force of infection locality (site MNJ.10) for example. See supplement for full data. **Figure 3C**: Time until infection reaches a target prevalence versus the force of infection (FOI). A threshold of 10% prevalence is chosen as an example, corresponding to the WHO-defined upper limit for a low malaria burden setting. Baseline observed FOIs for March for the ten sampling localities in Mananjary district are increased (to the left) or decreased (to the right) by the indicated percentage for each site to capture outcomes across a wide range of uncertainty in estimates of the FOI. Scenarios with a decreased FOI show the potential impact on return times of reductions in FOI associated with future prevention or treatment interventions that may reduce malaria transmission. For reference, the dotted horizontal line shows 90 days (approx. 3 months), with the area under the horizontal line (outlined in red) corresponding to scenarios where an interruption in access to prevention and treatment results in infection exceeding the target threshold. See supplement for results for all age groups.

#### 2.2 Incidence rebound results are robust to variation in the force of infection

Our initial estimates of FOI indicate that prevalence rebounds rapidly when drug-based treatment and prevention coverage ceases (e.g., reaching 10% for school-aged children within 8-92 days by site). Future changes to malaria transmission dynamics or uncertainty in initial estimation of FOI may result in variation in the rate of rebound. For example, undercounting infected hosts due to imperfect RDT sensitivity, or multiple infections in the same host, will result in an underestimation of FOI (**Figure S5**). To account for this, we evaluate a range of FOI scenarios (**Figure 3C**). Increasing FOI results in prevalence reaching the threshold prevalence level sooner (e.g., for school aged children, reaching 10% prevalence 3-31 days sooner for a 50% increase in FOI), demonstrating that rapid rebounds in prevalence are robust to RDT sensitivity or other sources of underestimation. Alternatively, FOI could be decreased by reductions in the intensity of transmission achieved by intervention prior to the disruption event, or improvements in access to routine clinical care. However, even in optimistic scenarios where earlier intervention substantially reduces transmission rates, prevalence rebounds are only slightly delayed at high FOI sites (e.g., by 8-9 days for a 50% decrease in FOI for sites MNJ.01 and MNJ.10). These data suggest chemoprevention-based intervention regimes with currently available drugs are of limited efficacy in maintaining a low prevalence of infection in the face of climate-driven disruptions for the FOI ranges likely in moderate to high transmission settings.

### 3. Vaccination under climate-driven disruption to control

#### 3.1 Modeling vaccination

An exciting shift for malaria control is the recent availability of anti-malarial vaccines. Vaccines have a substantially longer duration of protection (i.e., > 10 months (*52*, *53*)) than currently used chemoprophylaxis drugs and longer than the time interval relevant for seasonal, climate-driven disruptions such as tropical cyclones (i.e., where the the vast majority of storms occur within a 3-4 month window). . The recently authorized vaccines have shown modest efficacy against clinical disease: 50-56% for RTS,S (*52*) and 68-75% for R21 (*53*) . To evaluate the potential of vaccines to reduce malaria disease burden under climate disruptions, we quantify the reduction in burden under a range of scenarios of vaccine coverage, efficacy, and symptomatic rates of infection (see methods) (**Figure 4**). Symptomatic rates are obtained from our cohort study where we find 37.0-53.2% of infections among children are associated with observable symptoms (**Figure S6**), in line with previous estimates of asymptomatic infection rates in moderate to high transmission settings (without vaccination) (*54*).

**Figure 4:**
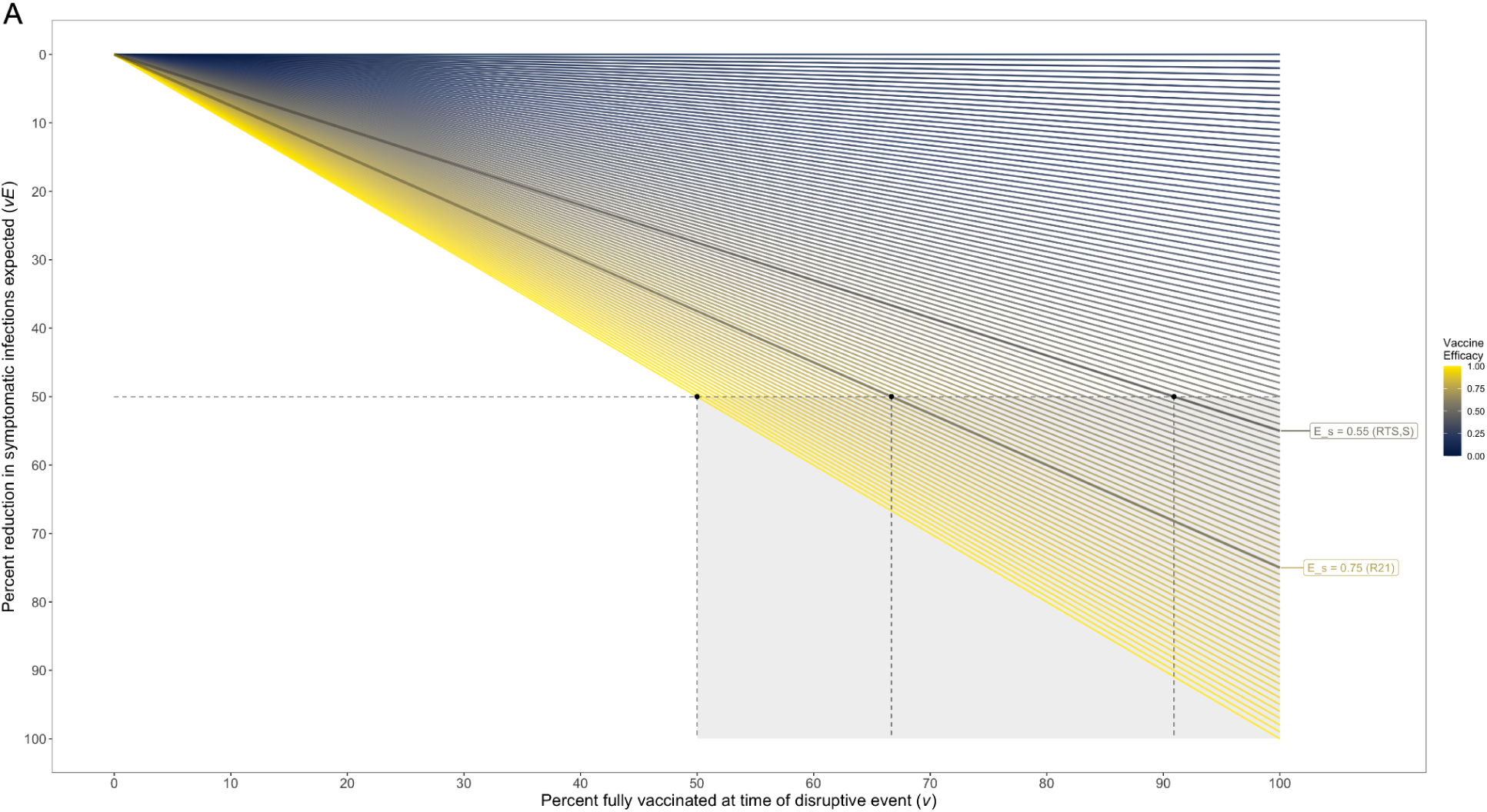
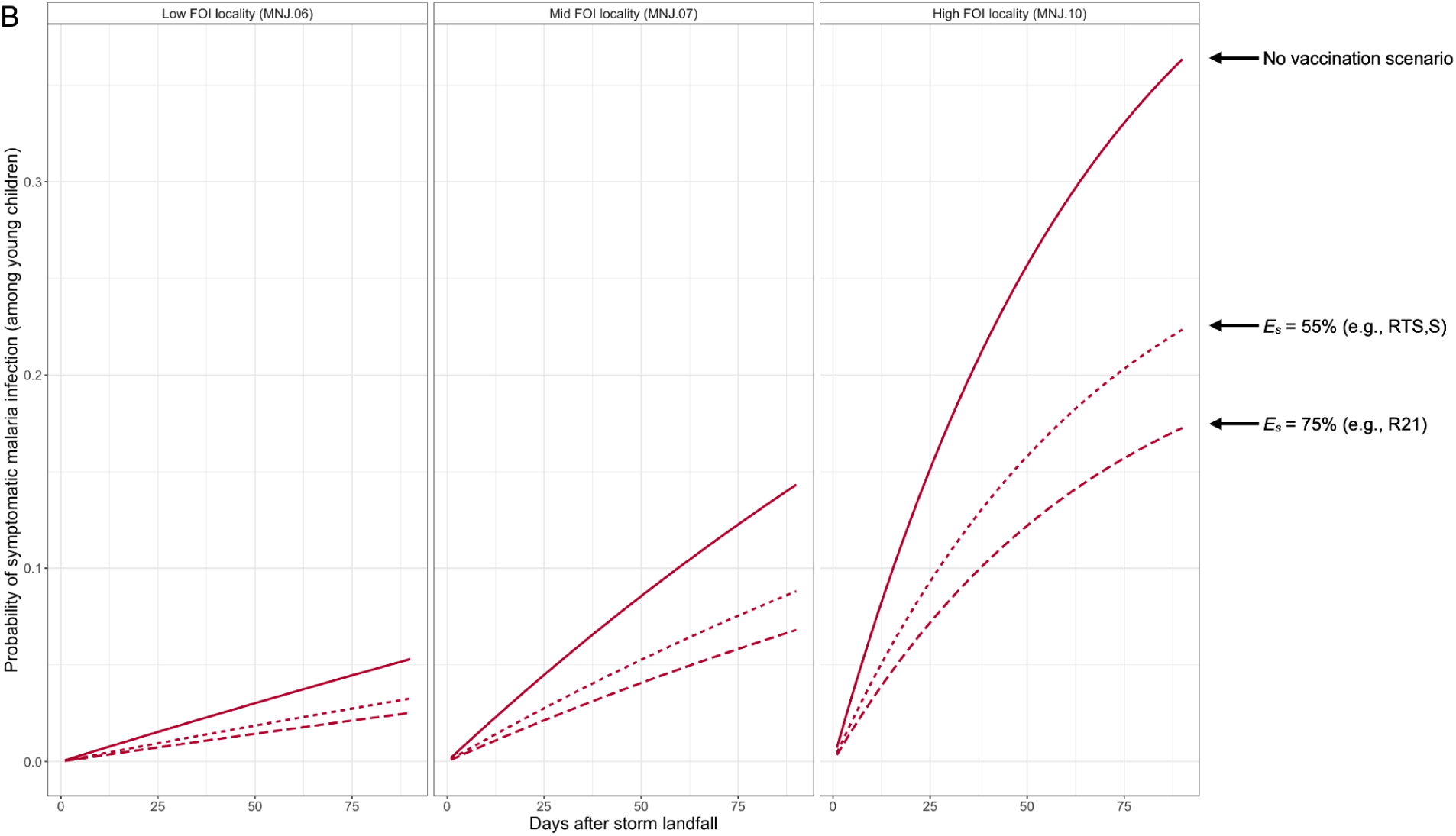
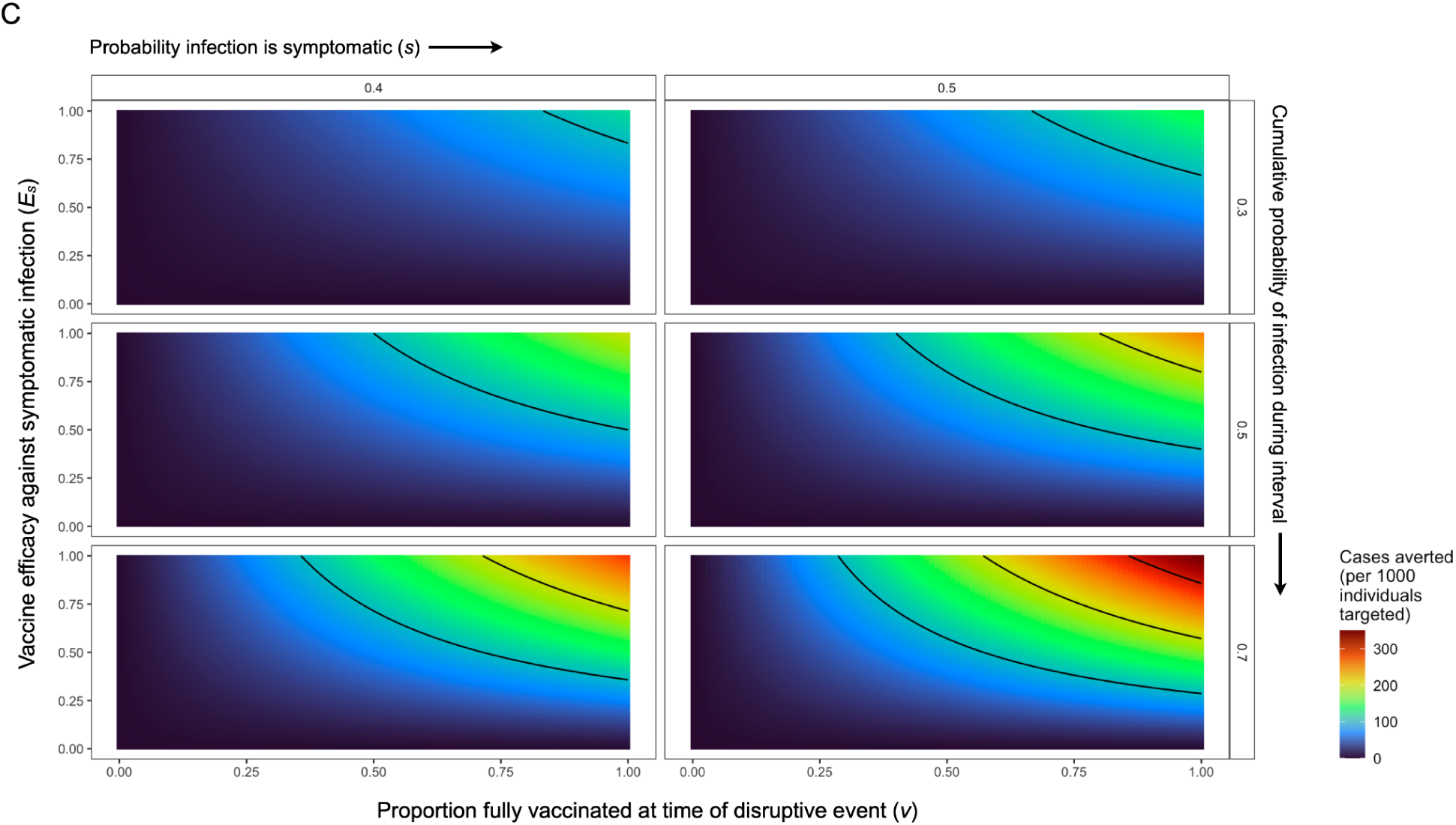
Symptomatic malaria infection probabilities in the presence of vaccination. **Figure 4A**: The percent reduction in the number of expected symptomatic infections across a range of vaccine efficacies. Estimates for the efficacy against clinical symptoms upon infection (Es) of the two currently available anti-malaria vaccines, RTS,S (*52*) and R21 (*53*), are shown for example. The shaded quadrant shows the scenarios with sufficient efficacy and coverage to observe a 50% reduction in symptomatic infections. See **Figure S7** for probability definitions and estimates accounting for partially vaccinated individuals. **Figure 4B**: Expected decrease in the cumulative probability of new malaria infections with vaccination, using the 90 day interval following Cyclone Freddy in 2023 for example. For illustration, estimates are shown for three localities: the highest observed force of infection (FOI) locality (MNJ.10), a mid FOI locality (MNJ.07), and the lowest observed FOI locality (MNJ.06). For full data, see Figure S7. Parameters here have been set to: proportion of infections symptomatic in unvaccinated individuals (s) = 50%, symptomatic vaccine coverage (v) = 70%, age = 5 years. Estimates of the efficacy (Es) for the RTS,S and R21 vaccines are as from panel A. **Figure 4C**: For the targeted subpopulation (i.e., children), the number of symptomatic malaria infections averted by vaccination with efficacy against symptoms *E_S_*. Rows show results for three levels of infection rates, expressed as the cumulative proportion of the population expected to acquire a new infection detectable by rapid diagnostic test (RDT) over the time interval considered. Columns show results for two values for the probability with which infections become symptomatic in the baseline population. Contour lines show the portion of the plot space where cases averted per 1,000 individuals targeted is above 100, 200, and 300 cases, respectively. See Figure S8 for the full range of parameters explored.

#### 3.2 Vaccination has the potential to reduce malaria burden following extreme weather events

When 70% of the targeted population has completed the full course for a vaccine with efficacy 68-75% (that reported for the R21 vaccine in phase 3 trials (*53*)) prior to the disruptive event, a 47.6-52.5% reduction in the expected proportion of symptomatic infections is expected (**Figure 4, Figure S7-8**). This level of vaccine coverage is plausible given the coverage (approx. 70%) reported for routine childhood immunization in the country (*55–57*). Existing estimates of vaccine efficacy are obtained on high background rates of access to and use of bednets and other vector control interventions (*58*). Estimates of bednet usage in Madagascar are 70-80% on the east coast of Madagascar (*59*) and at our study sites we estimate usage was 72.6-82.6% before and 69.4-76.4% after the two cyclone events (**Figure S9**). The number of cases averted is dependent on the infection rate and the symptomatic rate. Using the observed parameter values for the highest force of infection site (MNJ.10) and 50-70% coverage with the R21 vaccine as an example, we estimate, per 1000 children, a decrease from 135.1-193.5 symptomatic infections in the 2 months following a February cyclone to 64.1-101.4 symptomatic infections under vaccination (with 45.9-101.6 cases averted, respectively) (**Figure 4C**).

#### 3.3 Vaccination results are robust to modest efficacy and coverage

Vaccine completeness or timeliness may be imperfect for a fraction of the population, for example due to the timing of a child’s birth relative to planned vaccination dates or missed vaccination rounds due to population displacement. Additionally, efficacy may wane in subsequent years (*53*). To account for this variation in the degree of protection we explore a range of proportions of partially protected individuals (**Figure S7**). We also explore a range of symptomatic infection probabilities, including scenarios where symptomatic infection becomes more probable as infection derived immunity declines under lower incidence (**Figure S8**). We find vaccination can lead to a substantial reduction (i.e., approx. 50%) in the frequency of symptomatic malaria infection during disaster aftermath, results that are robust to even moderate vaccine efficacy and reasonable vaccination coverage (**Figure 4B-C**).

## Discussion

### 1. Temporal gap-filling as a potential use for malaria vaccination

Leveraging a dataset from Madagascar, we show that discontinuities in protection against malaria are a threat to efforts to reduce the burden of disease. Extreme weather events are a challenge to maintaining continuity. Climate change amplifies the risk of some extreme weather events (*60*), including tropical cyclones (*12*), with potentially devastating consequences relevant for health (*19*). The role of the disruptions likely to result from climate-driven disasters, and the best options for response, have been neglected in the study of climate change and malaria.

Vaccination, recently available for malaria, has a protective effect sufficient in duration to persist through disruptive events, such as cyclones and their aftermath. To date, assessments of the cost-effectiveness of vaccination, and priority areas for vaccine deployment, do not account for the additional benefit of long lasting protection against disease in times of disruption. Our findings suggest a new strategic use for vaccines, the deployment of vaccination in areas vulnerable to periodic disruption of public health activities. For climate-driven disruptions, historical (decadal) patterns of cyclone frequency could be used to define priority areas, potentially updated by projections of shifts in the future spatial distribution of tropical cyclones (*15*, *61*). Over shorter time scales, improved forecasts for an area’s cyclone season (*62*) may improve planning, although the limits of prediction may reduce the feasibility of anticipatory deployment of malaria intervention along an individual storm’s trajectory.

Disruptions associated with conflict may be less predictable, but interventions in settings where cyclone exposure and malaria incidence is moderate to high (e.g., Yemen (*63*), Somalia (*64*)) may help reduce burden. Additionally, vaccines using new technology such as mRNA may improve upon efficacy against clinical disease observed for vaccines available so far (*65*).

### 2. Limitations: Diagnostic uncertainty from reliance on RDTs and moderate vaccine efficacy

Ideally, estimates of infection rate would be determined with highly sensitive diagnostics (e.g., PCR) (*54*, *66*). Here, as in many settings with limited access to advanced health infrastructure, diagnostic options were constrained to field-deployable, rapid diagnostic tests (RDTs). We developed a set of sensitivity analyses to allow inference under the expected diagnostic uncertainty, with general applicability to other settings reliant on RDTs for surveillance. While RDTs are likely to miss low parasitemia infections (*67*), the extent to which these contribute to clinical burden, the target for vaccination, is unclear (*41*, *68*). Additionally, as described above, undercounting these infections minimally alters the magnitude of timing of infection rebound relevant for disaster response.

As malaria control programs prepare to scale up interventions to meet ambitious targets in coming years, investment across multiple control options must be evaluated (*69*). While control programs work towards reducing the often large, often asymptomatic, infectious reservoir, populations will remain vulnerable during any interruptions in prevention or treatment coverage. Benefits from partially mitigating any temporal gaps in coverage present an important case for considering vaccination, even given current shortcomings. Limited vaccine efficacy and questions as to cost-effectiveness across varied settings (*70–72*), alongside their minimal transmission blocking activity and restricted age range for which data are available (e.g., children < 4 years) mean that vaccines are unlikely to be deployed alone. Efforts that combine prevention approaches with different kinetics, even though they may appear redundant during periods without disturbance, are likely needed to lessen the impact of extreme weather events. Additional investment in mitigation of conditions that increase exposure post-disaster are also likely to be valuable including reinforcement of vector control interventions such as bednets during periods of population displacement (*73–75*), strengthening health facilities to minimize damage (*76*, *77*), and creative drug distribution approaches for prophylaxis that avoid dependence on storm-vulnerable infrastructure (for example, for intermittent preventive treatment during pregnancy, IPTp).

### 3. The need for more research on attribution and disaggregating mechanisms of risk

While our study was necessarily confined to estimates of aggregate risk of infection following a climate event, a challenging but important task for future investigations will be to separate the proportion of risk attributable to background and seasonal trends from the direct and indirect impacts of an extreme weather event. Tropical cyclones are likely to alter the risk of infection through multiple mechanisms such as flooding, population displacement, and destruction of housing and clinics (*19*, *23*, *78*), that interact to modify mosquito vector abundance, host exposure to infection, and access to and use of prevention and treatment. Where overall magnitude of infection is high, as in our study area, this alone is sufficient to dictate the burden experienced during disaster aftermath and the likely response to control measure scenarios. More nuanced results titrating the role of specific extreme weather associated factors would require deriving expectations for non-cyclone impacted years (or locations), for example leveraging tools from causal inference. However, the scarcity of long-term data with direct estimates of malaria infection rates and vector dynamics, and the high frequency of cyclones, precluded such analysis here. Both cyclone seasons of the present study (2022 and 2023) featured major storms and, at the national scale, Madagascar has experienced 48 cyclones with intensity at or greater than a category 1 hurricane in the 44 years since 1980. Additionally, storms interact with multi-year changes in underlying drivers of malaria risk, such as periodic mass bednet campaigns, and any residual damage to infrastructure from previous years’ storms.

These complexities around identifying a storm’s possible impact on transmission potential do not undermine our critical finding: any decline in the continuity of access to healthcare and the deployment of control measures will increase population risk of infection. As a result, data on total risk of infection following a climate event, even when the fraction attributable to the event is uncertain or low, will be valuable for planning and should be a priority for future surveillance efforts.

### 4. Implications for disease control in an era of increasingly extreme weather

The overwhelming majority of global malaria cases and deaths come from a subset of African countries (e.g., 20 African countries contribute 86% of global cases) (*1*). The context of progress in control efforts in these high burden settings, the broad effects of climate change, new recommendations for expanded chemoprevention, and the advent of vaccination create a new environment for malaria control. Health system and infrastructural disruptions in these high burden areas could be a major consequence of climate change for malaria. Continuity in public health efforts will be decisive for global efforts to reduce the burden of this pathogen. Severe tropical cyclones, a source of likely disturbance, have marked seasonality, making the timing of their potential impact predictable and enabling anticipatory interventions, such as vaccination. Other forms of disturbance (e.g., civil conflict (*79*, *80*)) may be less predictable in their timing, but could benefit from similar analyses. In addition to malaria, other infectious diseases subject to control regimes reliant on the regular distribution of drugs or vaccination may be vulnerable to climate-driven disruption (*27*, *81*). This warrants increased attention on the interaction between climate and the continuity of public health access.. Vaccination may be a key mitigation measure against climate-driven and other disruptions in the future.

## Materials and Methods

### 1. Study area: Mananjary district, southeastern Madagascar

#### 1.1 Malaria epidemiology context

Located in southeastern Madagascar, the Mananjary district (region Vatovavy) is a coastal district with perennial malaria transmission. The district extends approximately 60 km to the north and south of the district capital, the city of Mananjary (-21.22° S, 48.35° E) where the only hospitals in the district are located. As of 2018, 90.1% of the population of the district lives in rural *kaomina* (“communes”) (Institut National de la Statistique Madagascar).

From Malaria Indicator Surveys, malaria prevalence among children 5 years or younger in the east coast of Madagascar averaged 9-18% from 2011 to 2021 (*59*, *82–84*), placing the district in the WHO defined moderate transmission stratum. In terms of seasonality (**Figure S10**), higher prevalence was estimated for December to May from re-analyses of health facility data (*85*, *86*), with a peak around April (*87*). Few recent epidemiological studies with active sampling are available from the region, but prevalence varied from 3-46% by locality in 2017 from cross-sectional prevalence surveys (*44*, *88*).

#### 1.2. Background access to prevention, diagnosis, and treatment in Mananjary district

At the time of study, the primary prevention activity in the district was periodic mass bednet distribution campaigns (most recently in mid-2021). We observed a low frequency of diagnostics and treatment use outside that provided by our study (Figure S11). At the baseline sample (July-October 2021), only 35 of the 822 RDT positive individuals reported performing a diagnostic test in the previous 2 weeks (17 at the nearest public health facility, 2 at the Mananjary district hospital, and 16 at a private healthcare provider). Likewise, only 23 of the 822 RDT positive individuals reported taking anti-malaria medication (165 reported taking medication other than anti-malaria medication such as paracetamol, ibuprofen, or other medicines), indicating self-treatment is rare.

### 2. Extreme weather event data

Historical data on the occurrence of extreme weather events was sourced from the National Oceanic and Atmospheric Administration (NOAA) International Best Track Archive for Climate Stewardship (IBTrACS) data (accessed 31 July 2024) and EM-DAT from the Centre for Research on the Epidemiology of Disasters (CRED) (accessed 21 February 2024). See **Data S1** for the full list of weather events. Search criteria were all storms where the eye passed within a 60 nautical mile (approx. 111 km) buffer of a country’s coastline. This buffer was chosen because extreme rainfall and winds may extend across the diameter of a storm. Tropical cyclones classification followed the Saffir-Simpson scale used in the Atlantic basin. Mean monthly temperature and rainfall for the approximate midpoint of Mananjary district were sourced from WorldClim 2.1, accessed using the geodata package in R. All code and data files are available at ref. (*89*).

### 3. Longitudinal sampling

#### 3.1 Prospective cohort study

A stratified random sample of 500 households evenly distributed among 10 rural sampling clusters (communities or *tanana*) were recruited and enrolled during a baseline sample in July-October 2021. Informed consent or assent was obtained for study participants prior to enrollment. Enrolled households were followed longitudinally for 10 follow-up sampling time points. Clusters were rural (defined as more than 5km from the Mananjary district capital) and approximately 10 km apart on average.

Study procedures received approval from the Institutional Review Board (IRB) of the Harvard T.H. Chan School of Public Health (IRB#21-0111), Comité d’Ethique de la Recherche Biomédicale (CERBM) at Ministère de la Santé Publique de Madagascar (N°019/MSANP/SG/AMM/CERBM 2021), the Mananjary district health office, local government authorities, and community leaders.

Figure S1 shows the proportion of the cohort negative, positive, and unsampled by rapid diagnostic test (RDT) per time point. The participation rate, calculated as the percentage of enrolled individuals screened, ranged from 62.5-91.0% per time point. We consider individuals with visible bands for lactate dehydrogenase (LDH) or histidine rich protein 2 (HRP2) antigens as positive. The RDTs used were the AccessBio CareStart™ Pf/Pan RDT or Abbott Diagnostics Bioline™ Malaria Ag Pf/Pan RDT per manufacturer’s protocol. The duration between measurements (see below) indicates that persistent positives are likely to be rare (*90*).

#### 3.2 Screening for malaria and symptoms

All individuals in enrolled households were screened for malaria at approximately 2-month intervals (mean interval between samples = 57.4 days, median = 52 days, n = 20,718 total observations). On site diagnosis by RDT was performed at the time of screening and a blood sample was collected on a dried blood spot (DBS) for later molecular confirmation. Individuals, or a surrogate for younger children, also completed a questionnaire on bednet usage and symptoms adapted from refs. (*91*, *92*). Sensitivity analyses were performed to account for RDT sensitivity and specificity (**Figure S5**).

Individuals positive for malaria by RDT were offered treatment by an onsite physician with artemisinin-based combination therapy (ACT) using the standard first line treatment for Madagascar (artesunate amodiaquine, AS+AQ). To observe treatment adherence, at minimum the first dose of the three-day course of treatment was observed directly by the research team. Symptomatic individuals or individuals requesting a consultation were offered a consultation with an onsite physician.

#### 3.3. Accounting for RDT sensitivity and specificity

Due to imperfect sensitivity and specificity for RDTs, false positive and false negative RDT results may bias inference of infection rates. From a previous study, an estimated 97% of infections in the east coast region of Madagascar are by *Plasmodium falciparum* and 88% of RDT positive infections were confirmed by molecular follow-up (*93*). In studies of diagnostic performance, RDT sensitivity and specificity was estimated to be approximately 80-90% (*94*, *95*).

We use a binomial sampling approach to explore the effect of diagnostic error. We simulate ‘false negatives’ by drawing from a binomial distribution with probability 1 − *sensitivity* such that a fraction of RDT negatives become positive. Likewise, we simulate ‘false positives’ by drawing from a binomial distribution with probability equal to the *specificity* such that a fraction of RDT positives become negative. **Figure S5** shows simulations over a range of sensitivity and specificity values, where we repeat the force of infection estimation methods (described in the following section) for 500 replicates. These results illustrate minimal bias across plausible ranges of sensitivity and specificity. When the proportion of positive tests is small, accounting for possible false negatives results in an increase in the expected proportion infected. For sites with a high proportion of positive tests (i.e., approximately 50%), false negatives and false positives are similarly likely such that the expected proportion infected is minimally changed.

### 4. Estimating the force of infection

The interval before infection probability reaches a given threshold will hinge on target prevalence and the rate at which individuals become reinfected. This is typically characterized by a parameter λ, the force of infection, which defines the rate at which susceptible individuals become infected. The catalytic model (*96*) then defines the probability of evading infection up until time *t* as:

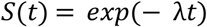

echoing results from survival analysis, and the probability of having been infected by time *t* (or cumulative density function of infection) is then the complement of this:

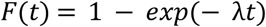

For malaria, infected individuals are susceptible to additional infection while currently infected (termed superinfection) and polyclonal infections indicative of superinfection have been reported from Madagascar (*93*, *97*). Given the shortness of the time interval considered here (e.g., <2 months), multiple infection events for an individual (super infection) are likely to be rare (i.e. < 1%) when the overall probability of infection per interval is low (i.e. < 10%); for sites or age groups where the infection rate is estimated to be high, we can adjust for possible underestimation for the force of infection given our inability to estimate superinfection by scaling it upwards (up to 100%) and evaluating the impact on our focal metric, i.e., infection rebound times (Figure 3C).

Because no evidence of resistance to ACT antimalarials has been reported to date from Madagascar, we assume the clearance rate of asexual and sexual stages of the parasite to be high. Given our direct observation of the early doses of anti-malarial treatment for infected individuals, we assume adherence to the treatment course is high and successful clearance of parasite infections following treatment. Thus for individuals positive at *t_i_*and *t_i+1_* we assume that a new infection has occurred over the time interval.

The force of infection may vary according to time-invariant covariates such as geographic site, and individual age (approximately time-invariant over the time-scales considered). We can fit these quantities using a generalized linear model fitted with a binomial likelihood using a complementary log-log link. Defining λ̂ as the estimated force of infection, and ti the length in days of time interval i, the estimated probability of being infected over interval i, is π_𝑖_ = (1 − 𝑒𝑥𝑝(− λ̂𝑡_𝑖_)). Applying the complementary log-log transform to this term yields η_i_ = 𝑙𝑜𝑔(− 𝑙𝑜𝑔 1 − π_i_) which expands to η_𝑖_ = 𝑙𝑜𝑔(λ̂) + 𝑙𝑜𝑔(_𝑡_) . Thus, fitting a model with η_𝑖_ = β𝑋 + log(ti) as the link function where β is a vector of coefficients, and 𝑋_𝑖_ is the corresponding design matrix of covariates for interval i, and log(ti) is fitted as an offset, we obtain a statistical expression that provides an estimate of the force of infection λ̂ as 𝑒𝑥𝑝 β𝑋_𝑖_ (*98*).

The set of covariates included in the design matrix 𝑋_𝑖_ can include indicators for factors (e.g., site) and smooth terms for continuous variables expected to fluctuate non-linearly (e.g., age). To include seasonal fluctuations in the force of infection, we must adjust for the fact that they are time-varying by introducing an additional covariate reflecting the number of days each individual at each measurement interval was exposed to different seasons of transmission (e.g., days in each month). Finally, we include a random effect of household to reflect the fact that individuals from some households were consistently infected over each time interval while those from other households were not. The final expression for the force of infection is then provided by:

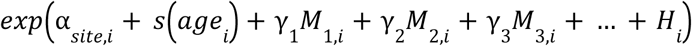

where the parameter α takes a different value for each of the ten study sites; 𝑠(𝑎𝑔𝑒_𝑖_) indicates a smoothed pattern across age, 𝑀_𝑗,𝑖_ reflects the number of days an individual was exposed to the 𝑗^th^ month of the year over interval i (directly obtained from the data) and the parameter γ_𝑗_ converts this into month specific effects on the force of infection over that interval, and 𝐻_𝑖_ corresponds to the random effect reflecting the household of individual 𝑖.

The statistical framework does not account for all uncertainties within the data. Underestimates of the FOI could emerge via i) false negatives returned by RDTs, which might also be more likely in low parasitemia infections in individuals having acquired some immunity, potentially particularly affecting FOI estimates for older individuals; ii) true positives reflecting superinfection, e.g., individuals likely to acquire additional infections when already infected, noting that some of this variability in infection risk may be accounted for by the household random effect. As our focus is on infection events, coinfection, i.e., simultaneous infection by multiple malaria genotypes from a single mosquito bite should not bias our results. Conversely, overestimates of the FOI could emerge in the presence of false positives, e.g., if RDTs remained positive long after the infection is cleared, although persistent positives are likely to be rare for our sampling interval (approx. 57 days on average) (*90*). Biases in our estimates emerging from these possible uncertainties are likely to fall well within the scope of the full spectrum of variance in the FOI that we explore.

### 5. Exploring the consequences of disruptions during intervention rollout

To explore the deployment of new mass chemoprevention and chemotherapy interventions, we simulate the early phases of these interventions, when the program is building towards, but has not yet, interrupted transmission. As a result, ongoing transmission results in the reaccumulation of infections, at a rate determined by the force of infection, following each intervention round until transmission is interrupted (i.e., force of infection approaches 0). We first investigate the return time, which we define as the maximum time interval between intervention rounds for a given force of infection where malaria prevalence is maintained below a chosen target level in the targeted population (see supplementary methods for full details).

The required program return time Δ_𝑐_ necessary to maintain prevalence below a chosen threshold 𝑧 is defined by:

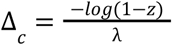

The estimated force of infection captures the rate at which individuals become infected, and interventions occur at intervals of Δ days, and clear 𝑝*_t_* percent of infections. Conservatively, we assume clearance for treated individuals to be 100% such that treatment successfully reduces infectiousness and treated individuals do not participate in transmission. This is supported by the lack of evidence of resistance to ACTs in Madagascar to date and evidence ACTs clear gametocytes (*99*). Subsequent to this intervention, mosquito vectors infected with parasites prior to the intervention or from contact with subsets of the population not covered by the initial round serve as a reservoir for infection. Thus ‘cleared’ hosts start to become infected again from continued exposure. The longer the interval Δ, the higher the prevalence of malaria reached within the target population.

We initially make no assumptions about feedbacks associated with transmission (relaxed in subsequent analyses), and additionally disregard recovery without treatment, since time intervals to recovery are typically long. Supporting this, published estimates of the distribution of duration of untreated *P. falciparum* infections (mean duration approximately 180 days (*41*, *42*)) indicate that the probability of recovery without treatment between our longitudinal samples (mean sampling interval 57 days) and during a disruption interval (e.g., 2 months) are very low.

### 6. Simulating additional control activities: Prophylaxis and vaccination

In addition to a primary intervention where a round of standard first line antimalarials (e.g., ACTs) is applied to clear existing infections, various interventions that prolong the duration of protection may be considered, from drug combinations with a longer prophylactic period to vaccination.

#### 6.1 Mass drug administration with longer lasting chemoprevention

Including a longer-lived antimalarial (e.g., sulphadoxine-pyrimethamine, SP, or dihydroartemisinin-piperaquine, DP) can prevent reinfection for the duration of effective prophylaxis. We define the mean duration of prophylaxis as *D*. We first consider a mass drug administration (MDA) scenario where all individuals, regardless of infection status, are given the ACT treatment paired with a prophylactic. When used to prevent reinfection for clinical cases this has been termed post-discharge malaria prophylaxis (PDMC). We also consider application in a mass test and treat (MTaT) activity where only individuals identified as positive for malaria infection receive the chemoprevention (see **Table S1** for definitions).

Because the drugs likely to be used in PDMC, MDA, and MTaT scenarios are likely the same as those for seasonal malaria chemoprevention (SMC) and intermittent preventive treatment (IPT), and thus have the same duration of protection, the return time for all can be modeled as *D* + Δ_𝑐_.

#### 6.2 Vaccination

Vaccination with the currently available antimalarial vaccines may provide a benefit in reducing clinical cases. We use estimates of the efficacy of antimalarial vaccines against clinical malaria reported from clinical trials for the RTS,S (approx. 55%) (*52*) and R21 vaccines (approx. 75%) (*58*, *100*).

In the simplest analysis, the expected number of clinically symptomatic cases over a short time interval (e.g., a 3-4 month cyclone season) results from the rate of infection, *λ*, giving the probability of being infected over an interval *p*(*I*) =1 − e^-λt^, and the probability of infection being symptomatic (*s*) or asymptomatic (1−*s*). Given no clinical data on efficacy against infection (*E_i_*) for currently approved vaccines, we conservatively assume *E_i_* = 0. For a proportion, *v*, of children fully vaccinated at the beginning of cyclone season, with an average vaccine of efficacy of *E_s_* (defined as reduction in probability of symptomatic malaria), we estimate the probability of symptomatic infection among children over an interval of length *t* following cessation of intervention due to a disruption as:

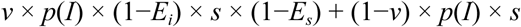

Similarly, scenarios where a fraction of the population, *v_p_*, is partially vaccinated can be considered. If the average protection against symptomatic infection for partially vaccinated children is *E_p_*(where 0 < *E_p_* < *E_s_*), the probability of symptomatic infection among children over an interval of length *t* can be approximated by:

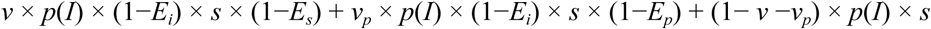

This conditional probability approach includes several assumptions: vaccine efficacy is similar across the age range targeted, vaccinated individuals are a random sample of the targeted population, no waning of protection, and that the fraction of the population targeted is insufficiently small to affect the population infectious reservoir. Consistent with these assumptions, vaccination to date has been deployed for small children only and efficacy seems to be maintained for more than a year after completion of the vaccination series (*53*).

## Data Availability

All data are available in the main text or the supplementary materials. Data files are available at a github repository: https://github.com/bennyrice/mananjary_cyclone_code

## Acknowledgements

We thank the CRS Multisectoral Malaria Project staff for support and the CRS Madagascar office and Mananjary malaria project staff for assistance with data collection: Dr. Virginie Andreas Nambinina Ralisoa, Dr. Elanirina Andrianoelivololona, Elodie Mialinjaka Rabarijaona, Tahiniony Tonie Ludjette Rakotozafy, Johnson Rakotohasimbola, Juloce Lidovique Razafindrakoto, Hermann Raelison Paratoaly, Aymardine Flashe Kantomananjara, Simon Razafindrasoja, Arisoa Beatrice Marie Sandrine Niry, and James Hazen. We thank the High Meadows Environmental Institute (HMEI) at Princeton University for support.

## Funding

CRS SCP4 Multisectoral Malaria Project (BLR, SM, MR)

The Falcon Award for Disease Elimination - The Climate Edit by the Global Institute for Disease Elimination (GLIDE) (BLR, MR)

The Ren Che Foundation and the AWS Impact Computing Project at the Harvard Data Science Initiative (HDSI) (CDG)

The High Meadows Environmental Institute (HMEI) at Princeton University (BLR, GAV, CJEM)

## Author contributions

Conceptualization: BLR, MR, JL, CDG, AW, BG, CJEM Methodology: BLR, AW, BG, CJEM

Investigation: BLR, ER, SM, MR, HR Visualization: BLR, BG, CJEM

Funding acquisition: BLR, MR, JL, CJEM, CDG Project administration: BLR, MR, JL

Writing – original draft: BLR, BG, CJEM

Writing – review & editing: BLR, ER, JL, GAV, AW, BG, CDG, CJEM

## Data and materials availability

All data are available in the main text or the supplementary materials. Code and date files are available at a github repository (*89*). Restrictions on human data: Only age category and not birthdate information is included in the publicly available data.

## Supplementary Materials List

Figures S1-S11

Tables S1-S2

References 101-103

Data files S1-S9

Data S1

Tropical cyclones and related humanitarian disasters in Madagascar since 1980 (xlsx)

Data S2

Locality information for study sites in Mananjary district, Madagascar including site codes, administrative division names, and coordinates of midpoints (xslx)

Data S3

Line list data of individual rapid diagnostic test (RDT) results and sample dates (csv)

Data S4

Tropical cyclone tracks data from IBTrACS (csv)

Data S5

Global estimates of average annual tropical cyclone exposure from Jing et al 2023 (*101*) (csv)

Data S6

Spatial estimates of *Plasmodium falciparum* incidence rate for 2020 from the Malaria Atlas Project (tif)

Data S7

National level estimated incidence of malaria from the World Health Organization (xlsx)

Data S8

Bednet usage questionnaire response data for the Mananjary cohort study (csv)

Data S9

Symptoms questionnaire response data for the Mananjary cohort study (csv)

---

**Figure S1.**
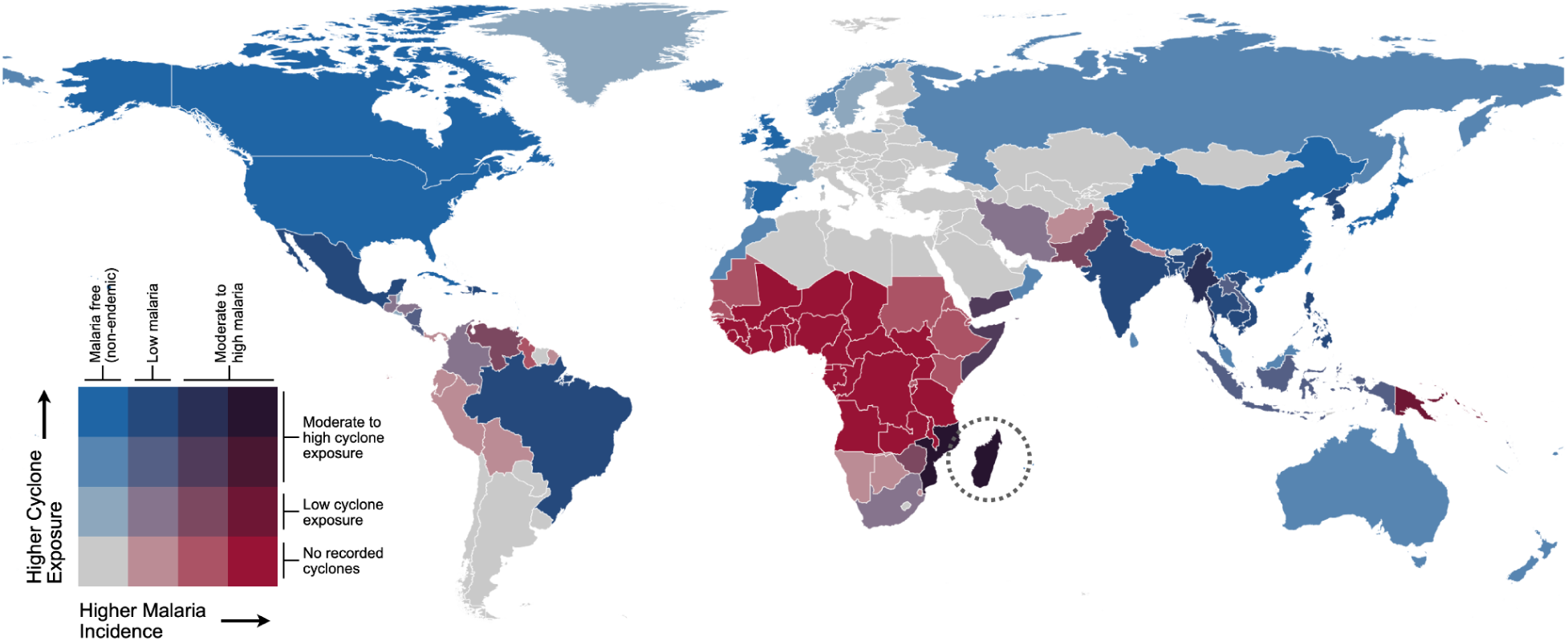
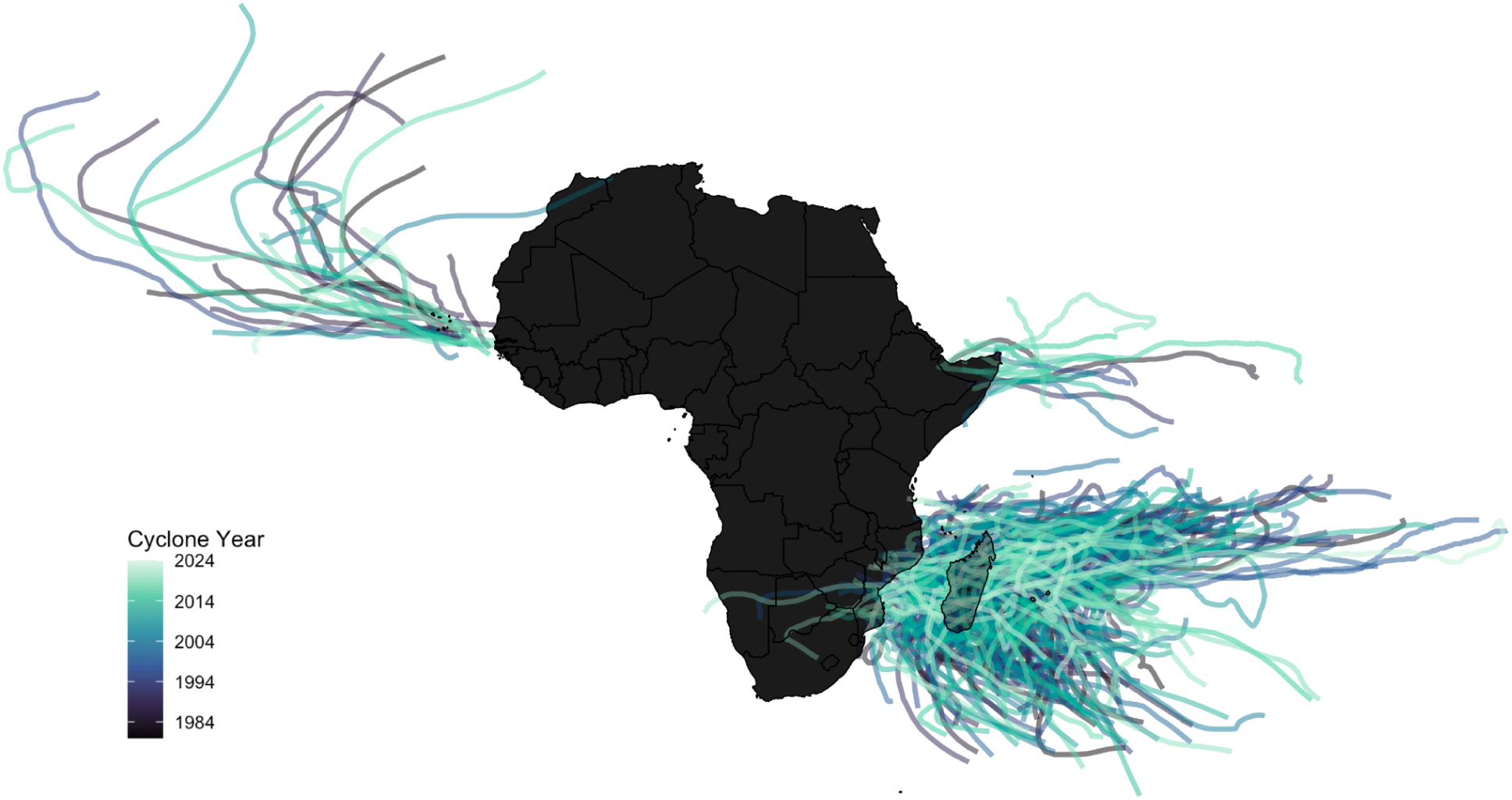
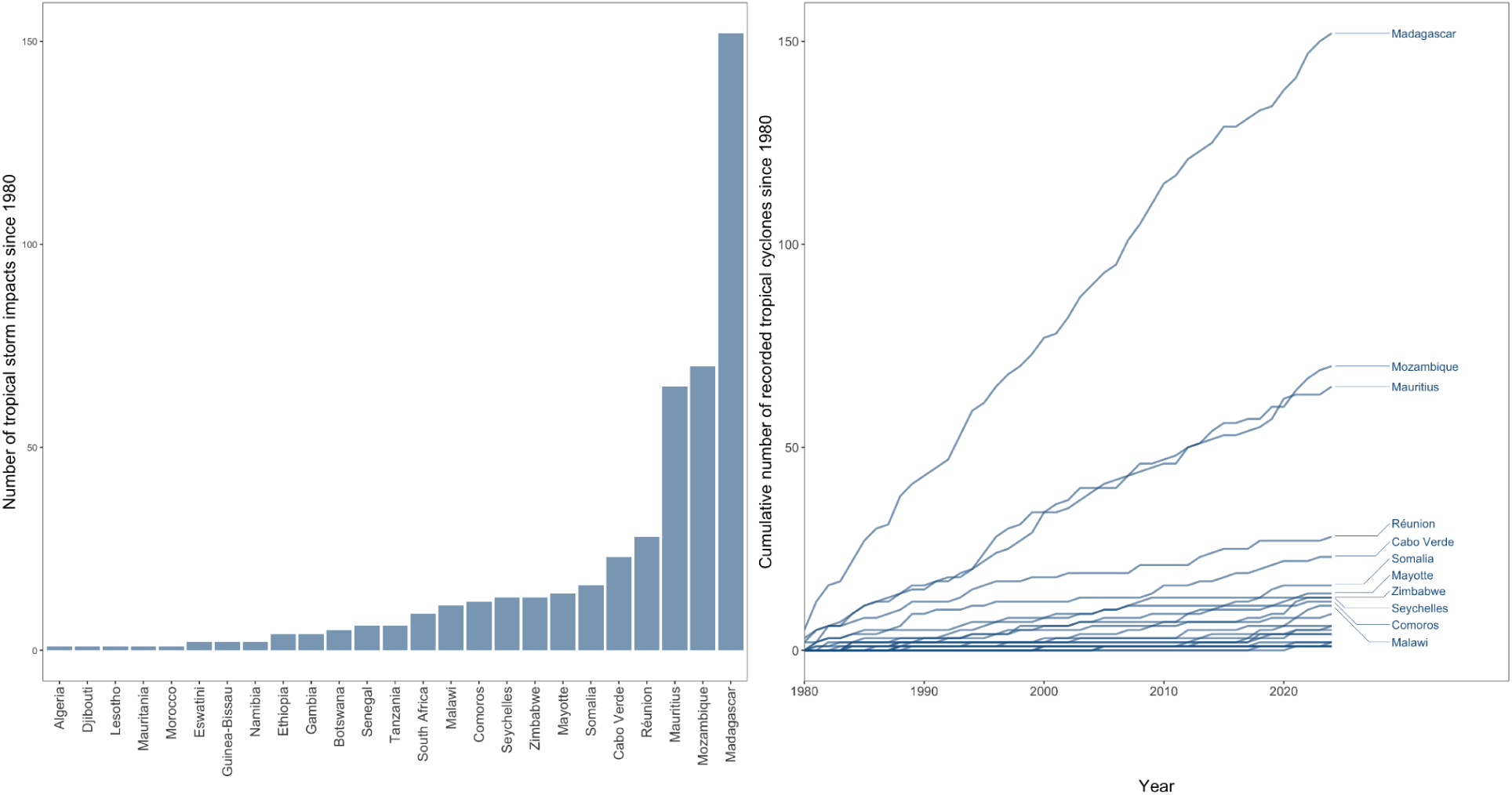
Tropical cyclone exposure and malaria burden extended data. **1A**: Malaria burden versus cyclone exposure at the national scale. Bivariate fill color (*102*) shows average annual person-days of exposure to tropical cyclones from 2002-2019 (*101*) vs reported national malaria incidence (*103*). Countries with no reported malaria cases or cyclone exposure are in gray. Madagascar circled for reference. **1B**: Tropical cyclone tracks in Africa colored by year of the storm (tropical cyclones since 1980 passing within a 60 nautical mile buffer of a country’s border from IBTrACS). **1C**: Tropical storm counts by African countries. Over 47.9% of impacts in Africa overall contributed by Madagascar and Mozambique (defined as the number of tropical cyclone tracks from IBTrACS since 1980 passing within a 60 nautical mile buffer of a country’s border).

**Figure S2.**
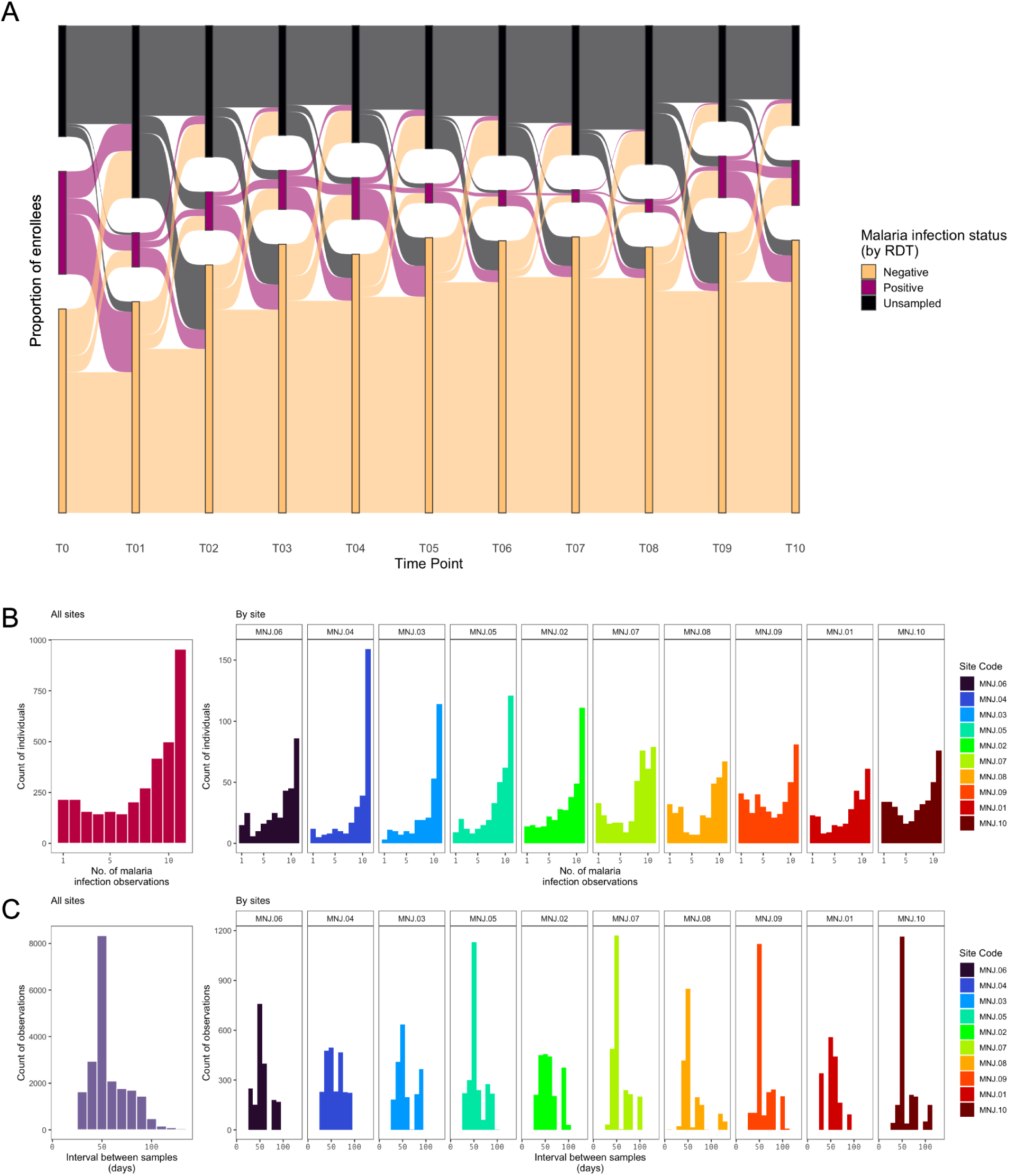
Sampling for the Mananjary malaria cohort study. **2A**: The proportion of enrolled individuals unsampled, negative, or positive by rapid diagnostic test (RDT) per time point from baseline (T0) to final sample (T10) (all sites combined). **2B**: Distribution of the number of observations per individual (site code colors and order follow Figure 2C). **2C**: Distribution of the interval in days between samples.

**Figure S3.**
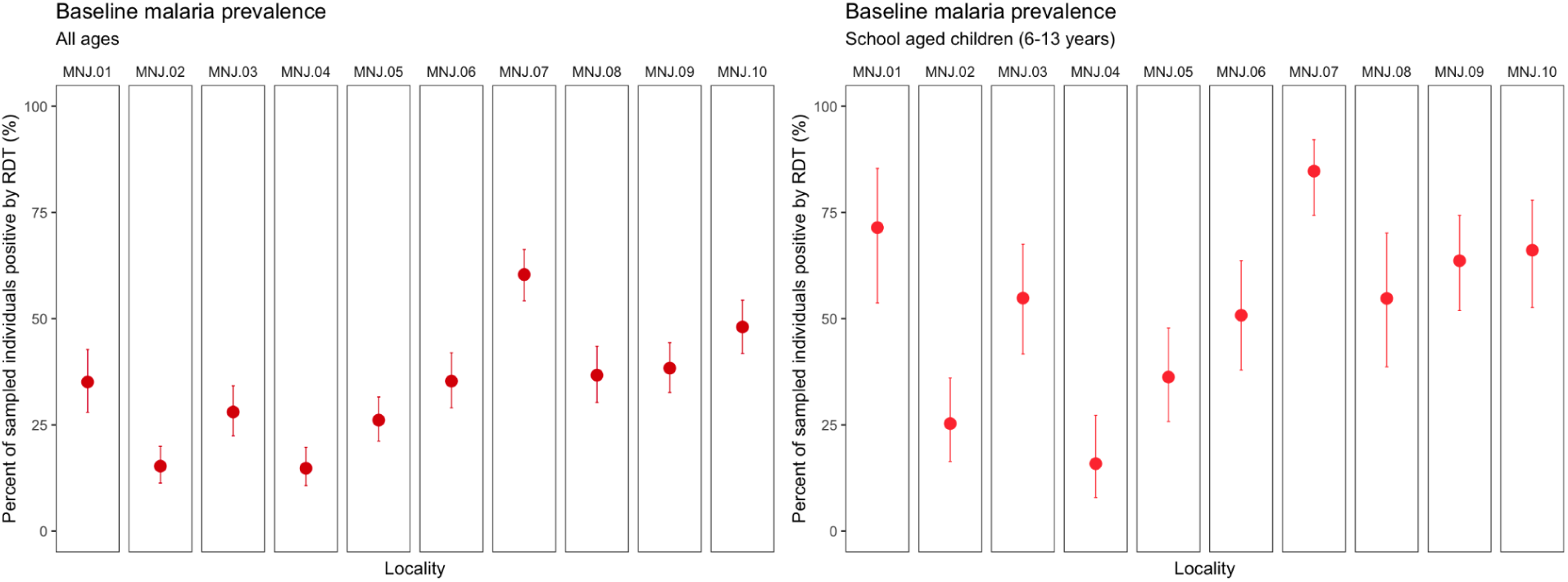
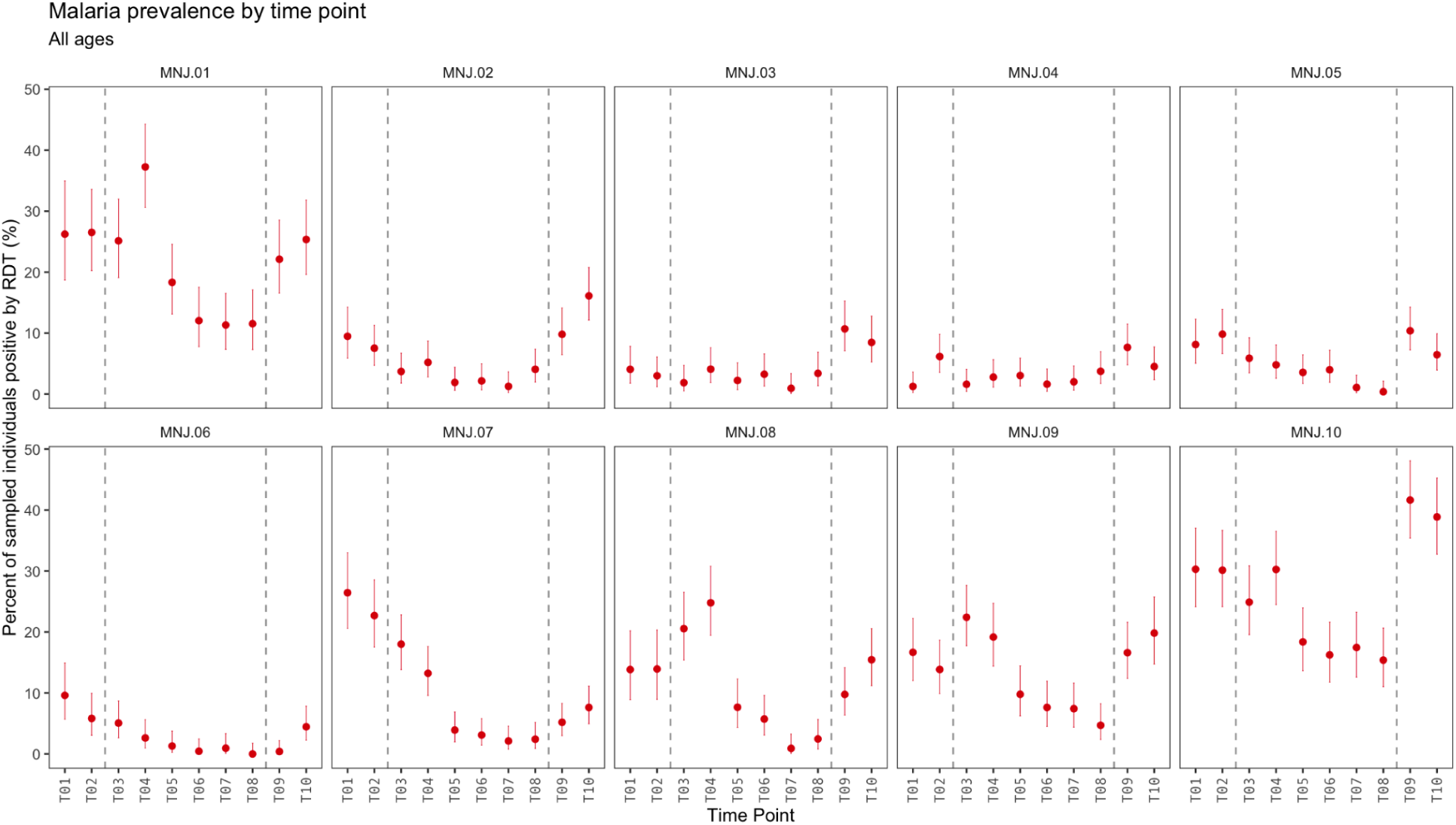
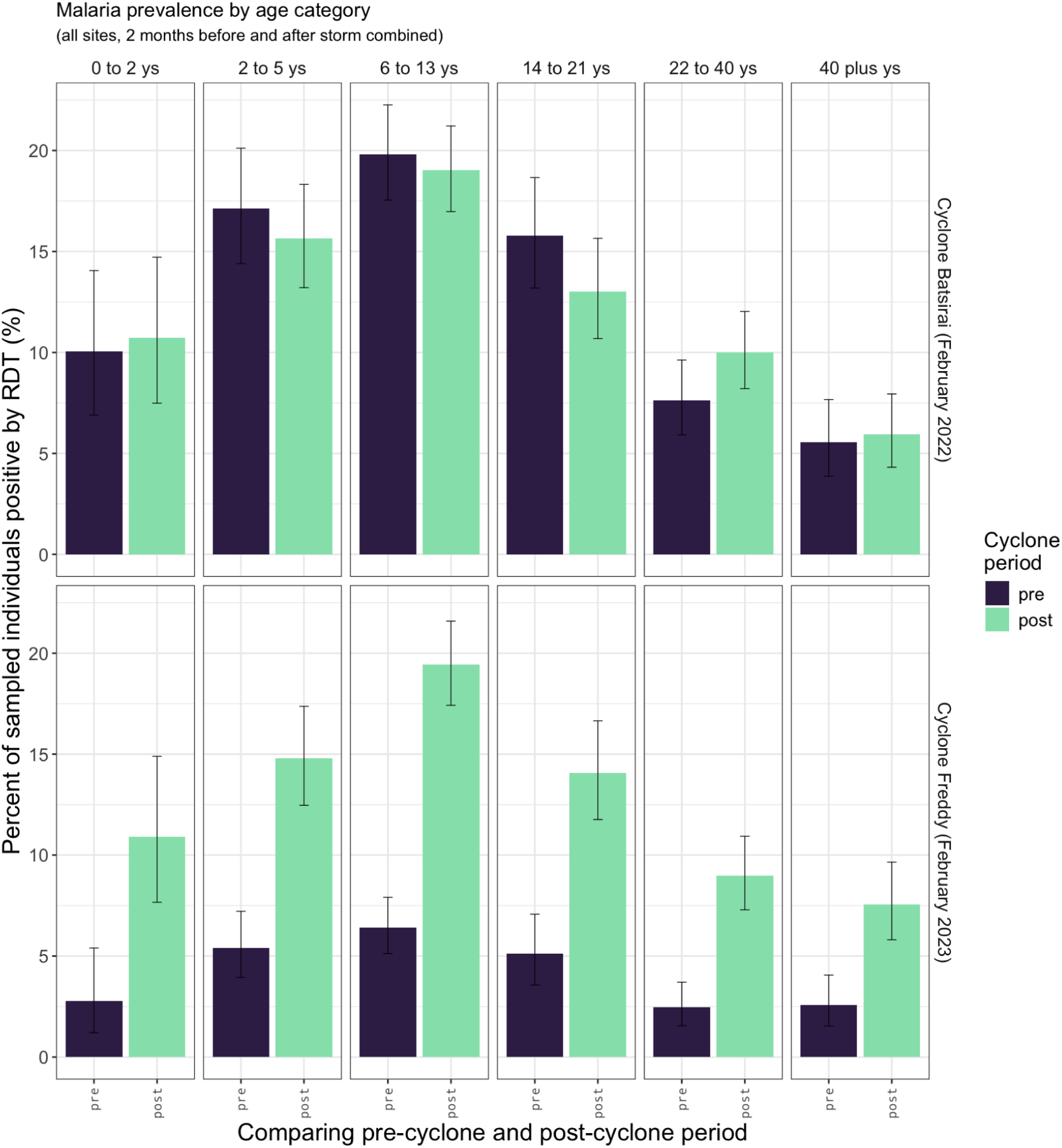
Malaria prevalence by time point and age. **3A**: Baseline prevalence of malaria infection (by rapid diagnostic test, RDT) for individuals enrolled in the Mananjary cohort study. Error bars show the 95% confidence intervals using the Clopper and Pearson (1934) method as implemented in the R binom.test function. **3B**: Prevalence by time point and locality. Vertical dashed lines show the timing of cyclones during the course of the study. Error bars show the 95% confidence intervals using the Clopper and Pearson (1934) method as implemented in the R binom.test function. **3C**: Age distribution of malaria infection by RDT in the two samples prior to and two samples following Cyclones Batsirai (February 2022) and Freddy (February 2023). Error bars show the 95% confidence intervals using the Clopper and Pearson (1934) method as implemented in the R binom.test function.

**Figure S4.**
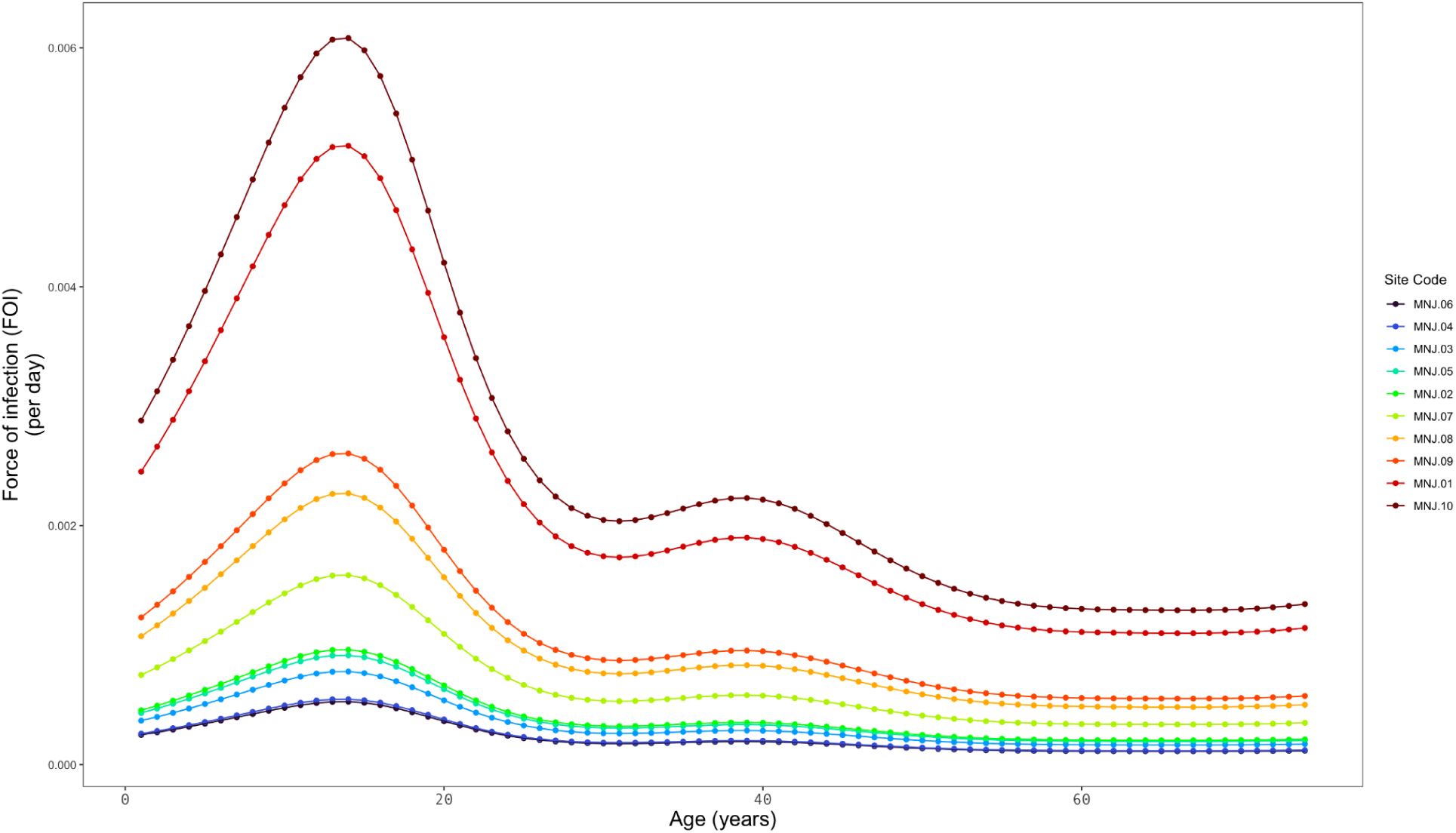
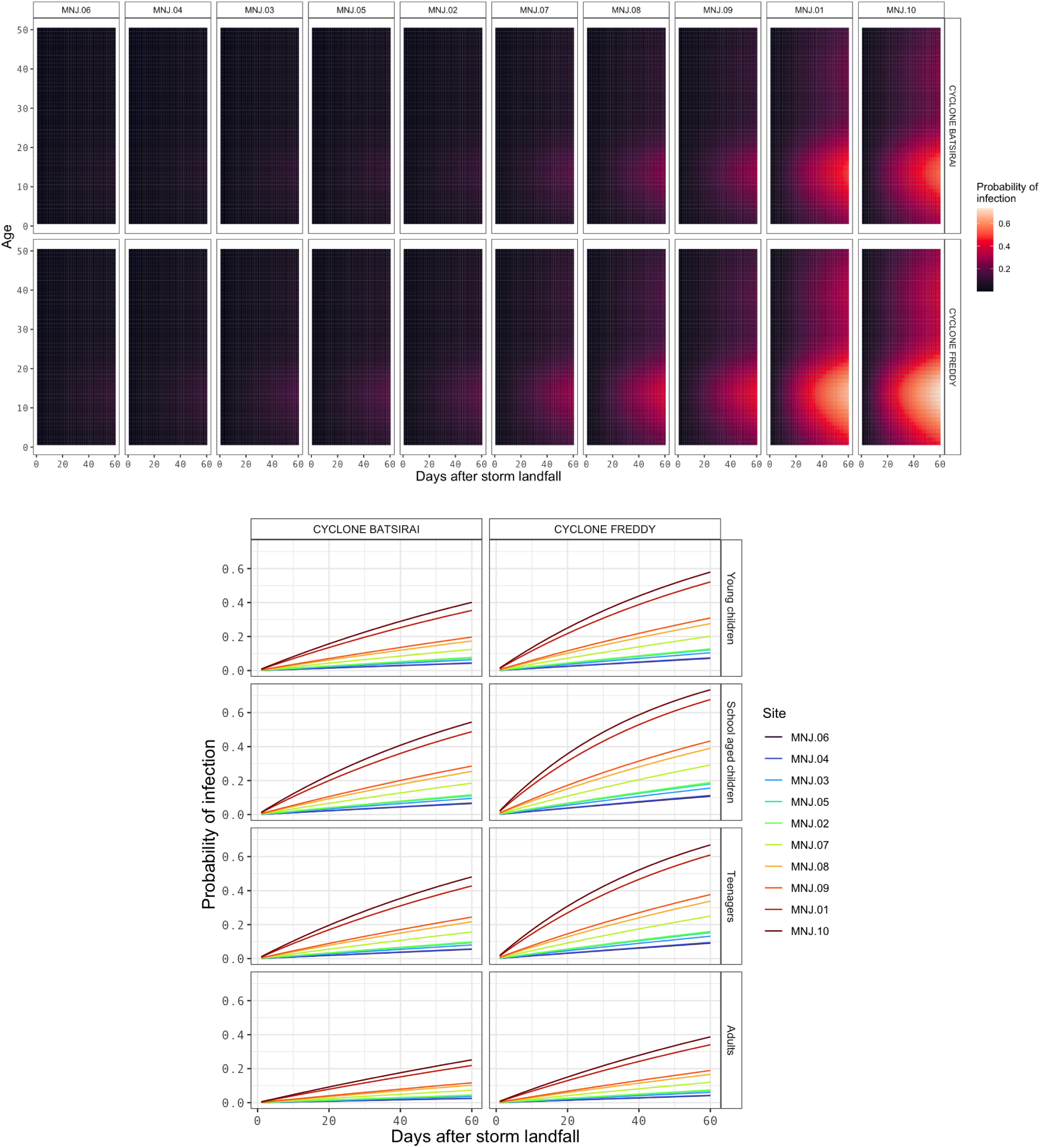
Force of infection (FOI) by age. **4A**: Observed variation in the force of infection by age for the 10 sample sites in Mananjary district, Madagascar (site code colors and order follow Figure 2C). Data shown for the month of January. **4B:** Estimated probability of a new malaria infection detectable by rapid diagnostic test (RDT) after Cyclone Batsirai (February 5, 2022) and Cyclone Freddy (February 21, 2023) landfall at study sites in Mananjary district, Madagascar. Sites ordered by infection rate. Age categories: young children = 0-5y, school-aged children = 6-13y, teenagers = 14-19y, adults = 20+y. Sites ordered by infection rate.

**Figure S5.**
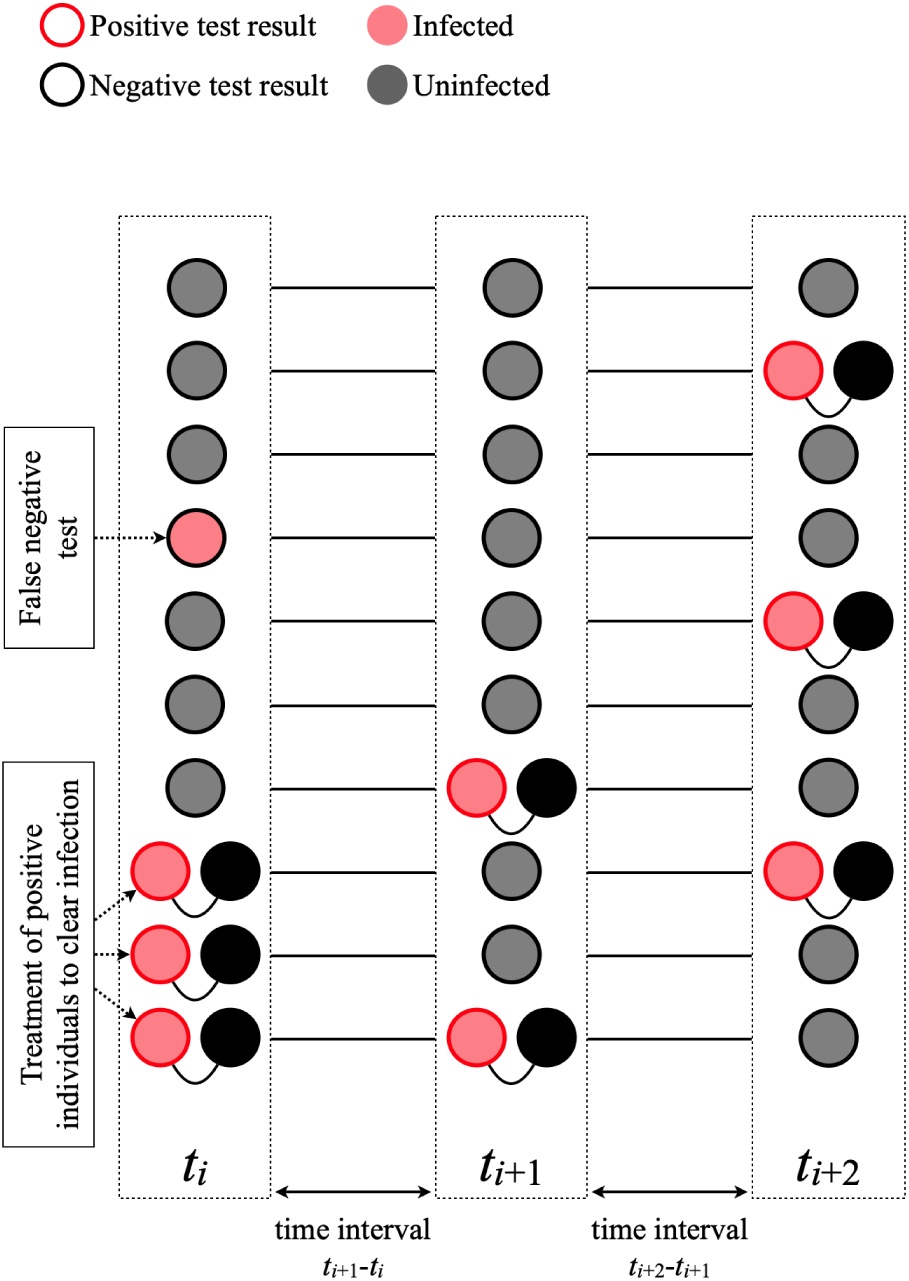
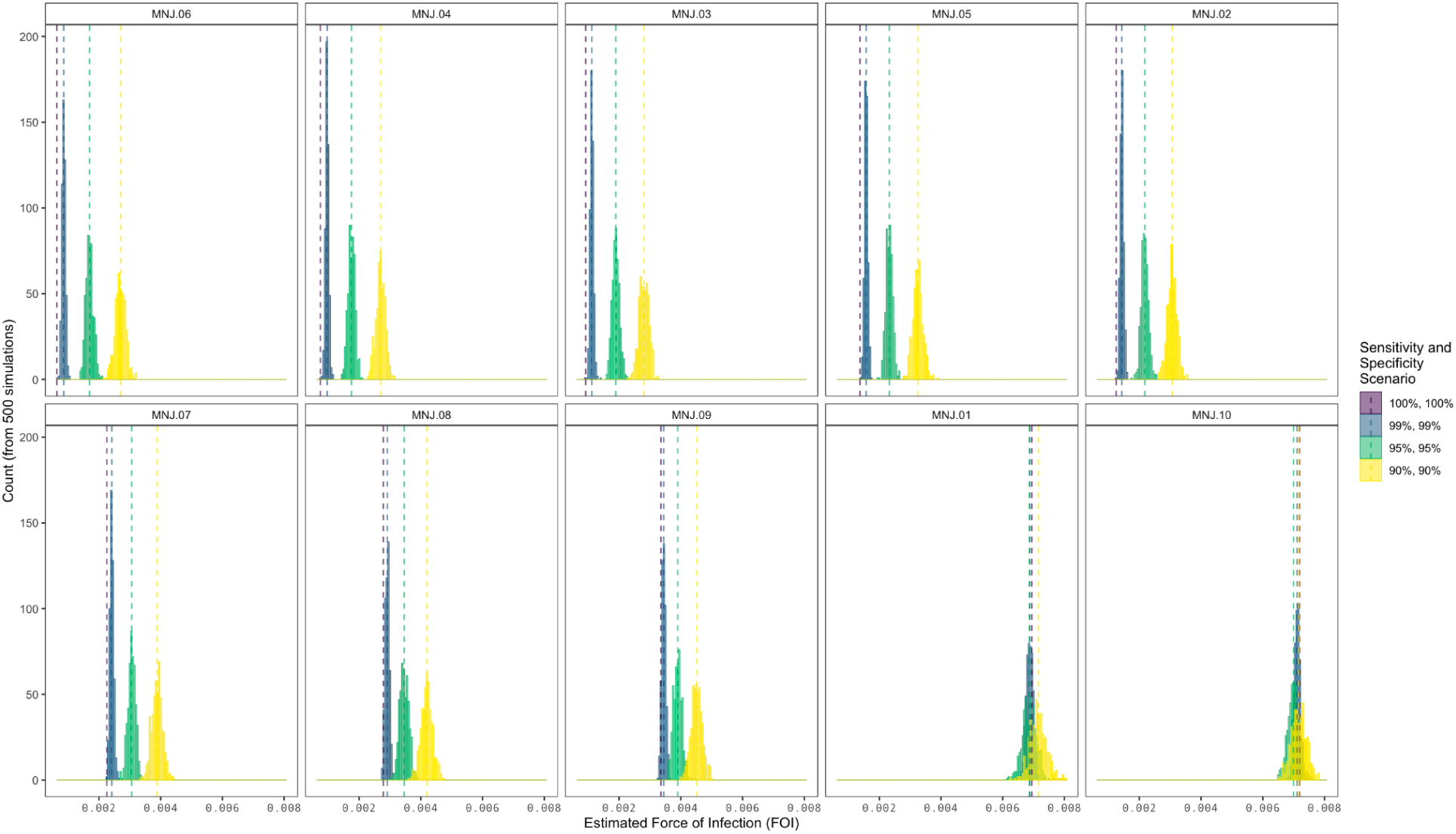
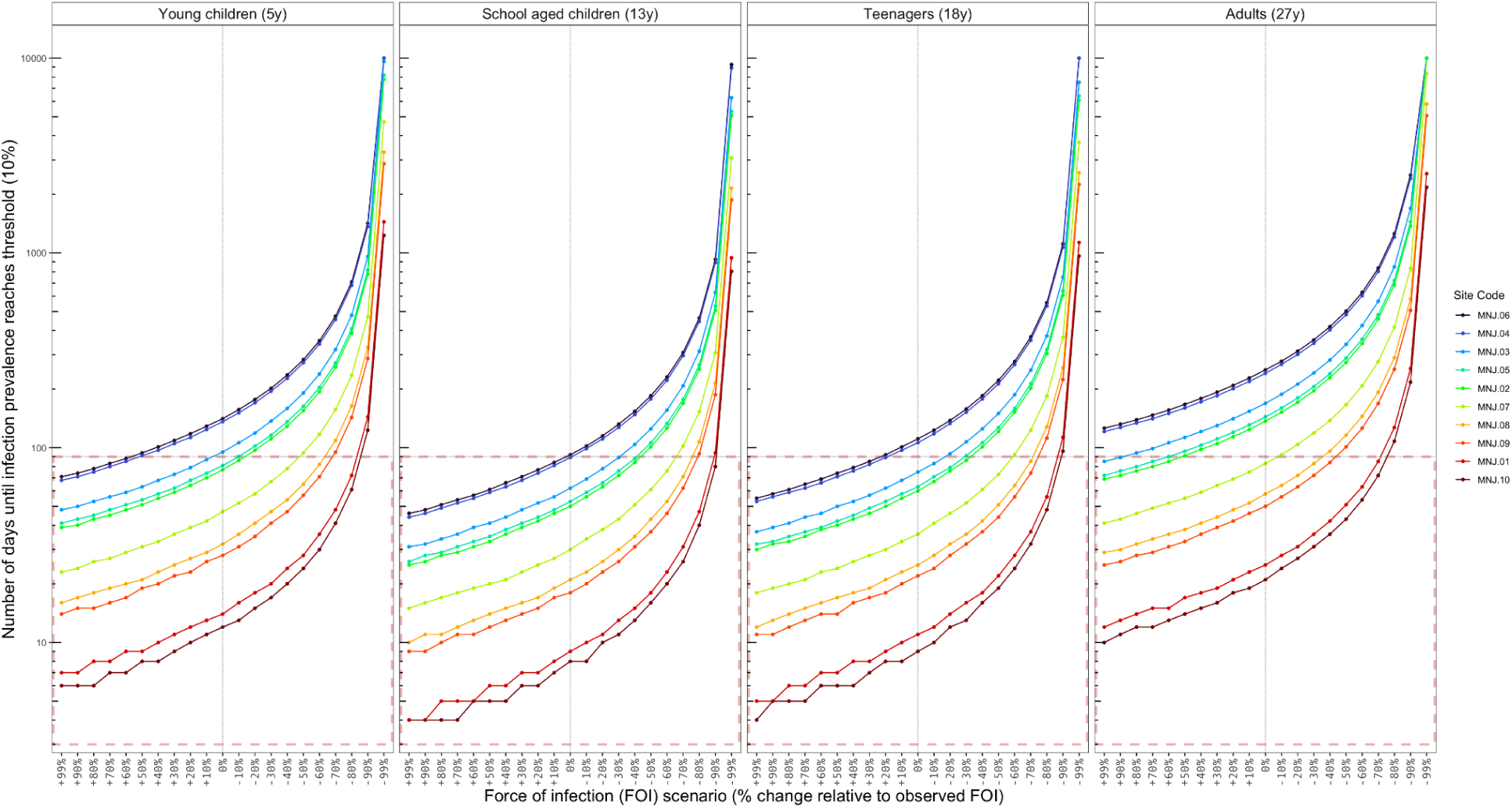
Sensitivity analyses for estimating the force of infection. **5A**: Schematic overview of sampling methods. Malaria rapid diagnostic test (RDT) positive individuals are treated with anti-malarials such that infection status at *t_i_*+1 can be assumed to be independent of status at *t_i_*. Some infections are uncounted due a false negative RDT result (shown as circles with black outline and red fill). Likewise, infections that clear before the next sample will not be observed (though this is likely rare as the average infection duration is greater than the average sampling interval). **5B**: Sensitivity analysis of RDT diagnosis. From 500 simulated trials, the force of infection (FOI) per day for a given sensitivity and specificity. Sites are ordered from lowest to highest observed rate of infection. Dashed vertical lines show the mean FOI from 500 simulations. For low infection sites (e.g., Sites MNJ.06 and MNJ.04, shown at top left), reduced sensitivity increases estimates of FOI (e.g., FOI increases from an initial estimate 6.3 × 10^-4^ to a mean estimate of 2.7 × 10^-3^ for sensitivity and specificity = 90%). This indicates for low infection rate sites, our estimates for the probability of infection over a time interval are conservative, with false negative rapid diagnostic tests contributing to a higher probability of infection. For high infection sites (e.g., Sites MNJ.01 and MNJ.10, shown at bottom right) estimates of FOI are relatively invariant across the range of sensitivity and specificity values explored. False negatives and false positives largely offset such that the initial FOI estimate 7.19 × 10^-3^ is similar to the mean estimate (7.16 × 10^-3^) from the 500 simulated trials with sensitivity and specificity at 90%. **5C**: Time until infection reaches a target prevalence versus the force of infection (FOI). A threshold of 10% prevalence is chosen as an example, corresponding to the WHO-defined upper limit for a low malaria burden setting. Baseline observed FOIs for March for the ten sampling localities in Mananjary district (shown with vertical dotted line) are increased or decreased by 0-99% for each site four four exemplar age groups. The dotted horizontal line shows 90 days (approx. 3 months), with the area under the horizontal line (outlined in red) corresponding to scenarios where an interruption in access to prevention results in infection exceeding the target threshold.

**Figure S6.**
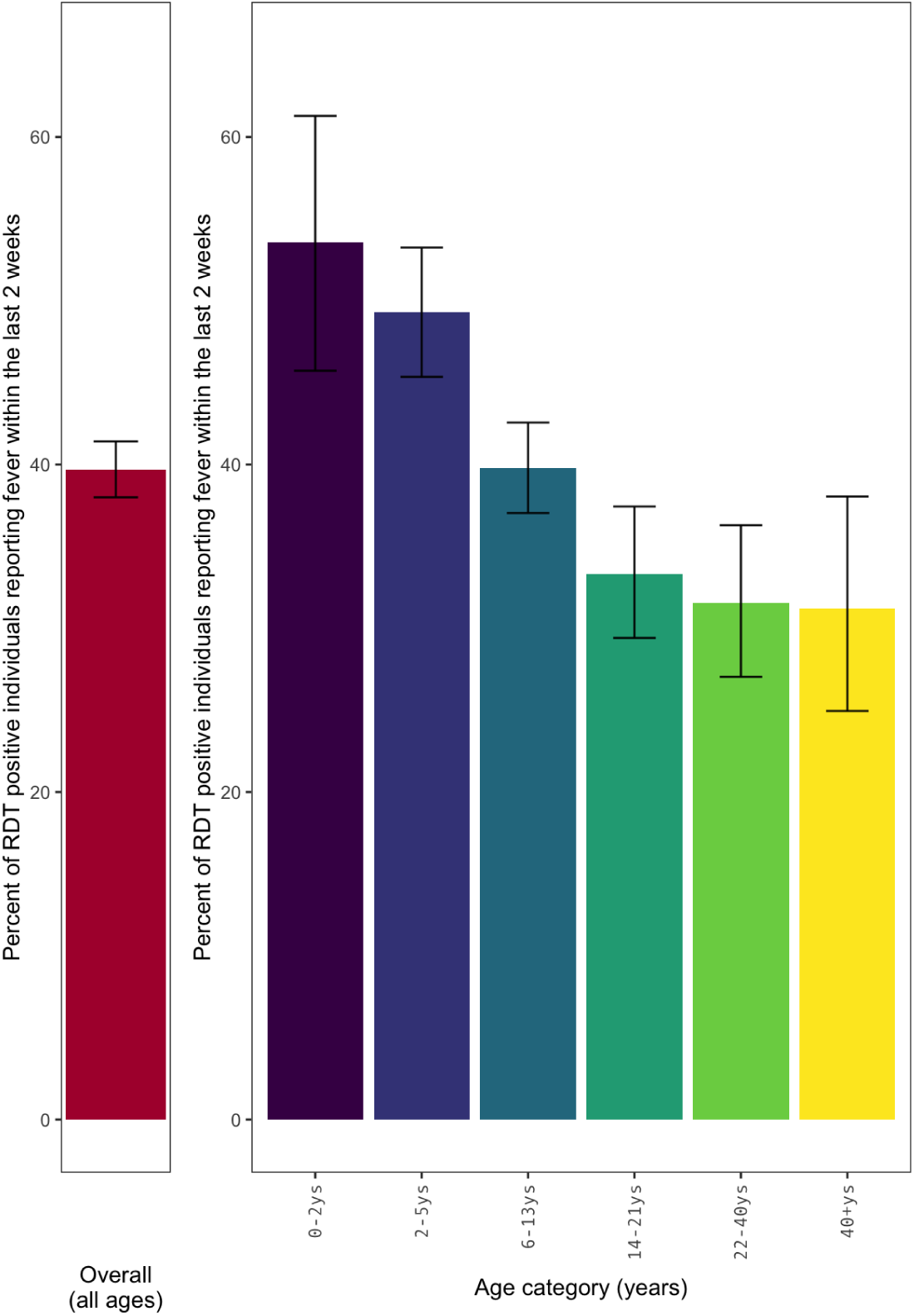
Symptomatic rates of infection by survey recall. The percent of rapid diagnostic test (RDT) positive individuals reporting any fever within the last two weeks of screening. Error bars show the 95% confidence intervals using the Clopper and Pearson (1934) method as implemented in the R binom.test function.

**Figure S7.**
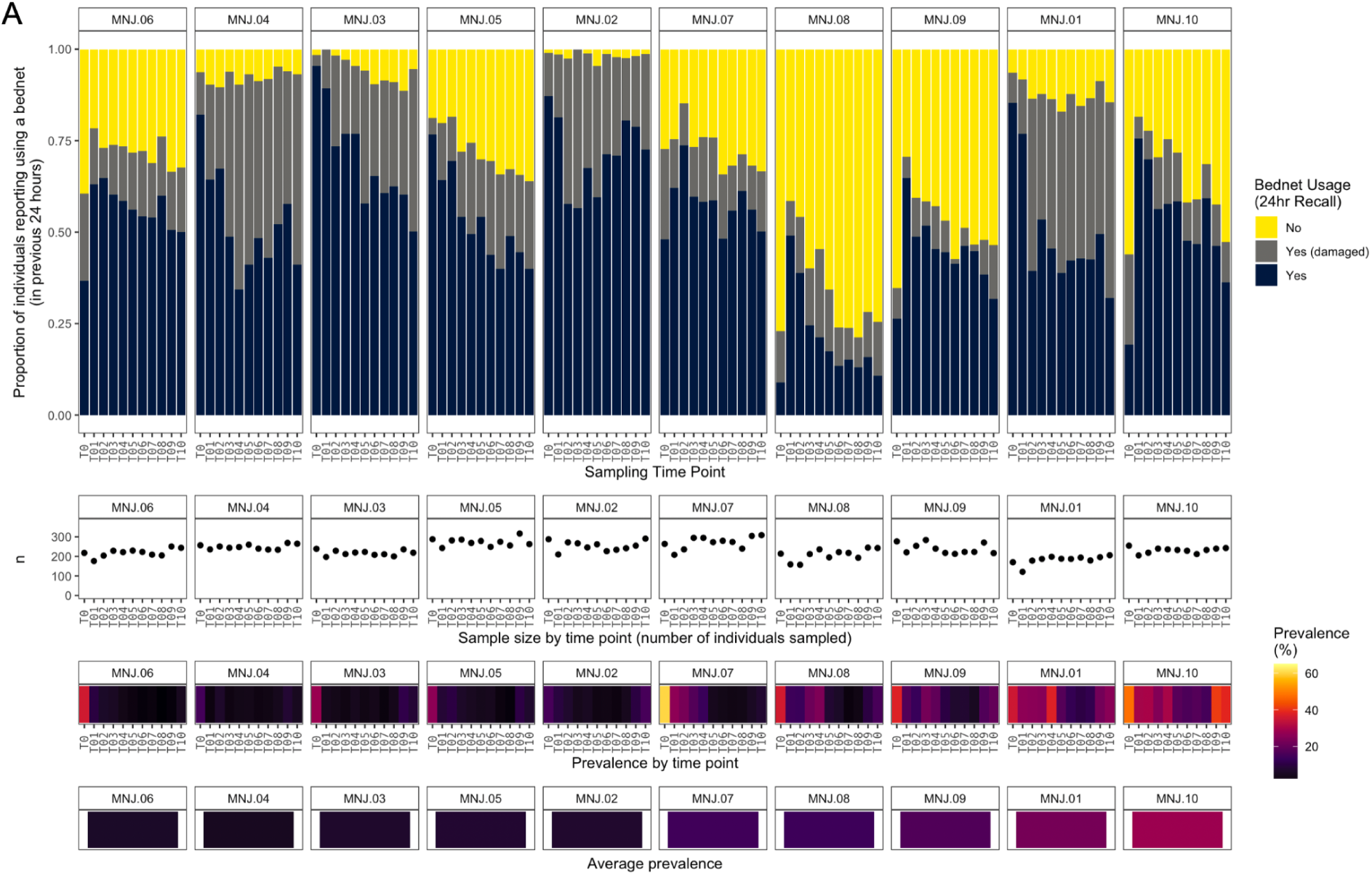
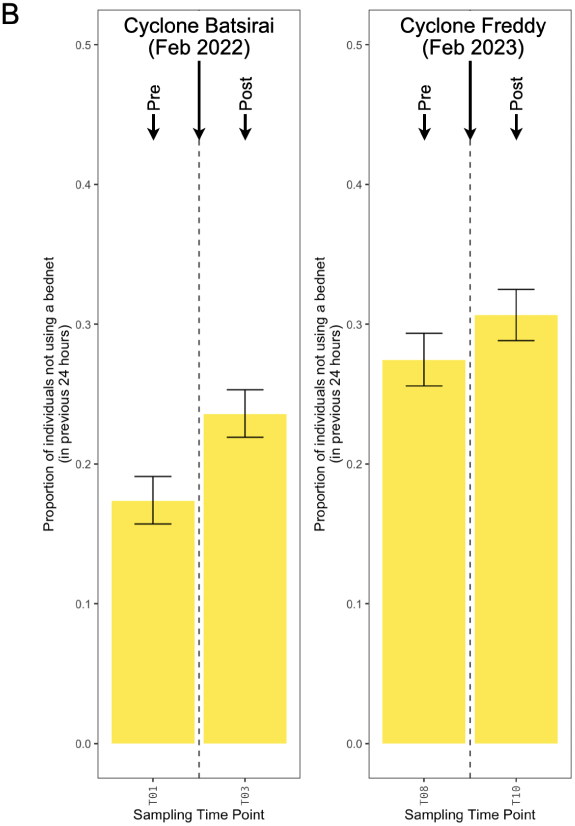
Bednet usage by survey recall. **7A:** The proportion of individuals reporting sleeping under a bednet in the last 24 hours and reporting bednet was damaged (due to holes, tears, or other damage). Sample size (n) and prevalence of malaria by rapid diagnostic test (RDT) for each site is shown below. Sites are ordered by average prevalence. Individuals tested by RDT have corresponding bednet usage recall data (range: 121-317 individuals per site per time point, mean: 234.6 individuals per site per time point). **7B**: The change in the proportion of individuals reporting not using a bednet for sampling time points before and after two cyclones. Error bars show the 95% confidence intervals using the Clopper and Pearson (1934) method as implemented in the R binom.test function.

**Figure S8.**
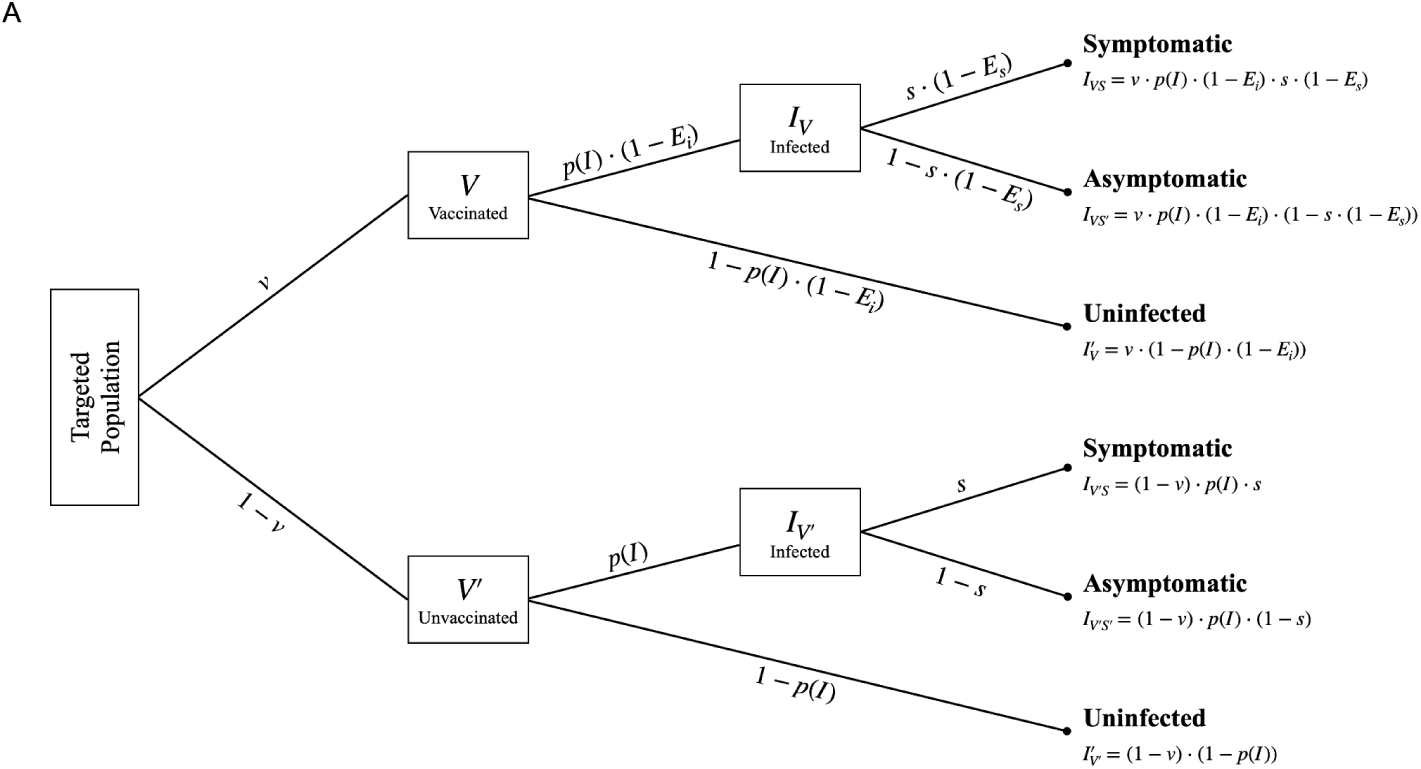
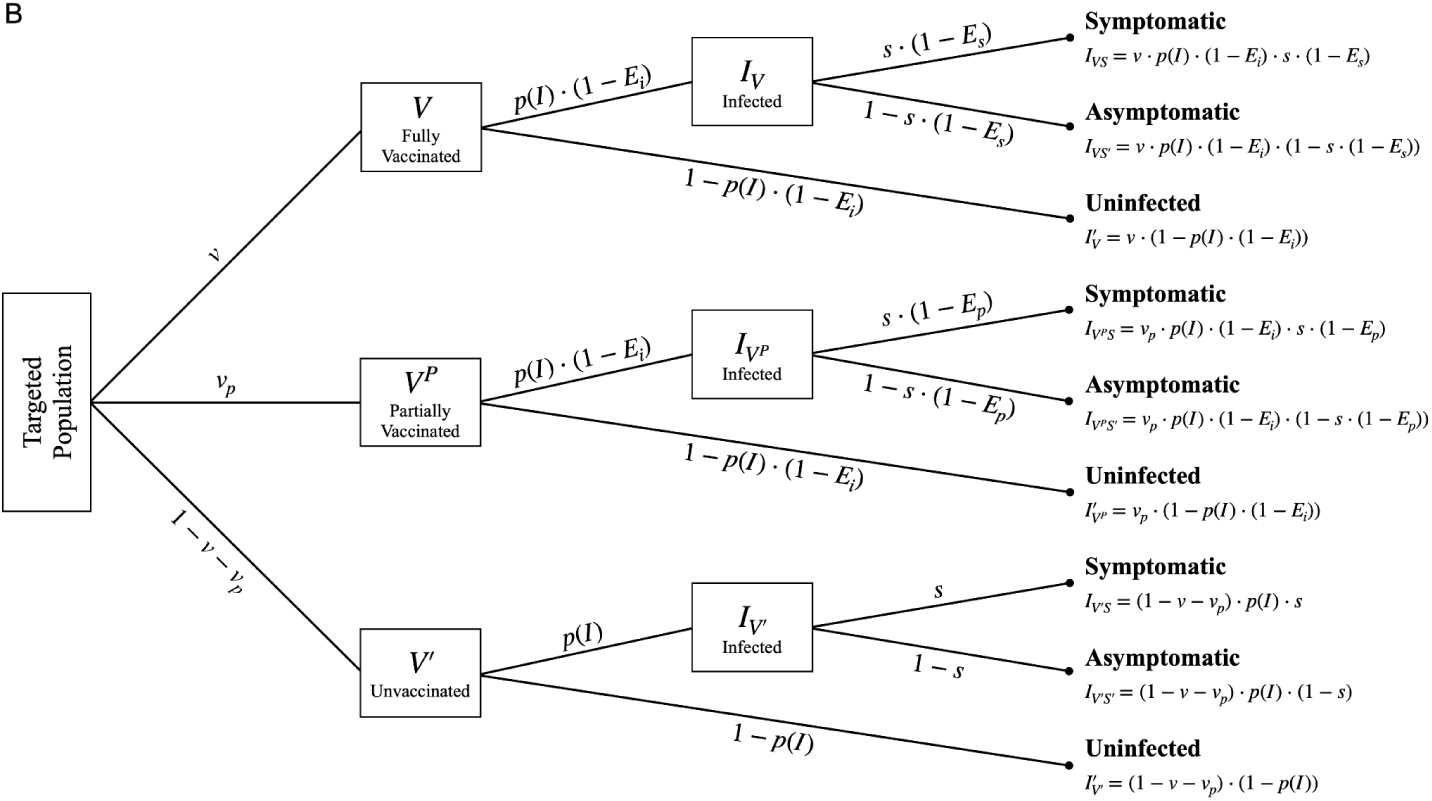
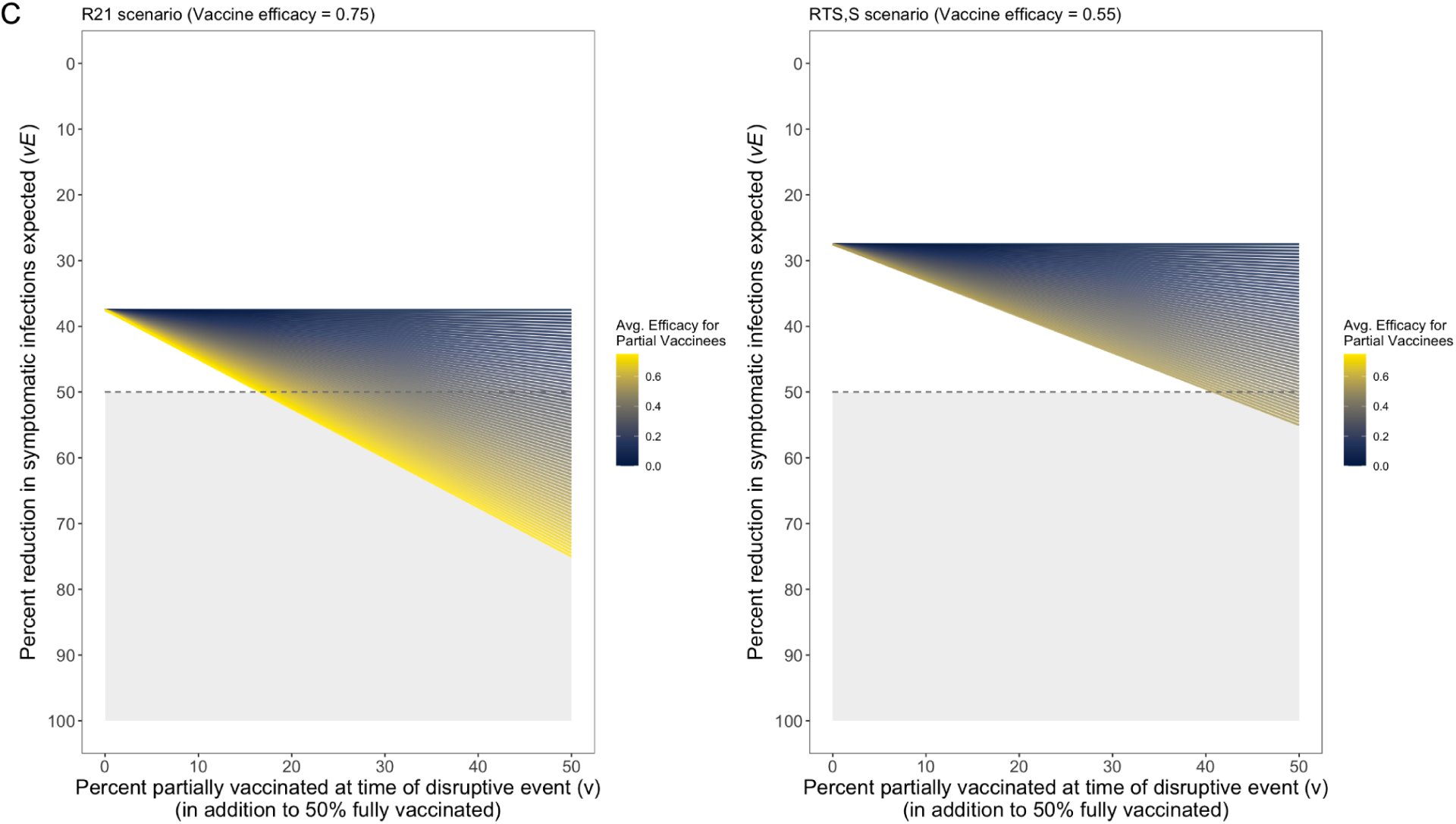
Simulating vaccination and partial vaccination. **8A**: Probability tree of the outcomes of malaria infection under vaccination. Parameters on branches specify the probability of either path at branching points. In the simplest analysis, the probability of symptomatic malaria infection for an age group over a time period *t* is determined by the probability of infection, *p*(*I*) = 1 − e^-*λt*^ where *λ* is the force of infection, the vaccination coverage, *v*, the probability of symptoms given infection, *s*, and any protective effect of vaccination, *E*. Vaccines may have efficacy against infection, *E_i_*, or against symptoms, *E_s_*. Note that when *E_i_* is small, as is thought to be the case for currently available vaccines, the probability of infection is similar for vaccinated, *V*, and unvaccinated, *V’*, individuals. When *E_s_* is large, vaccinated individuals are unlikely to have symptoms upon infection. **8B**: Probability tree of the outcomes of malaria infection under partial vaccination. The vaccinated branch can be separated further into partially and fully vaccinated sub-branches. Shown here assuming protection against infection, *E_i_*= 0, and protection against symptoms for fully vaccinated individuals *E_s_*is greater than the mean protection against symptoms for partially vaccinated individuals *E_p_*. **8C**: The percent reduction in the number of expected symptomatic infections across a range of partial vaccination rates with partial vaccine efficacy. Partial vaccination may result from failure to complete all doses of the vaccine series due to movement (e.g., displacement outside the area targeted for vaccination) or a lack of consistent access to vaccine delivery times or locations. Two example scenarios are shown where 50% of the targeted population is fully vaccinated and 0-50% of the population is partially vaccinated with protection varying from 0 to equaling the efficacy seen in fully vaccinated individuals (*E_s_*). The shaded area shows the scenarios with sufficient efficacy and coverage to observe a 50% reduction in symptomatic infections.

**Figure S9.**
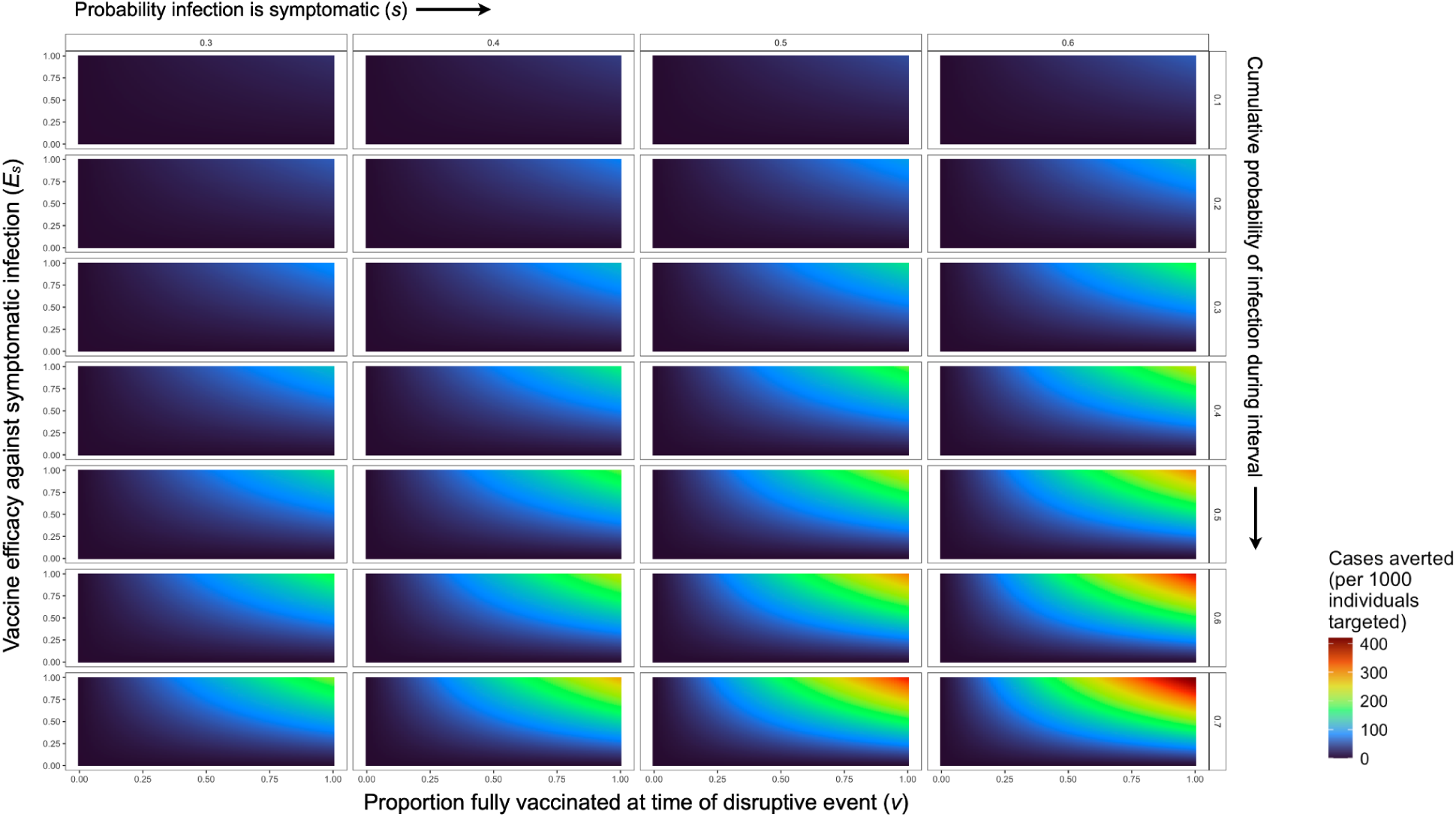
Sensitivity analyses for vaccination simulation. For the targeted subpopulation (i.e., children), the number of symptomatic malaria infections averted by vaccination with efficacy against symptoms *E_S_*. Rows show results for seven levels of infection rates, expressed as the cumulative proportion of the population expected to be infected over the time interval considered. Columns show results for four values for the probability with which infections become symptomatic.

**Figure S10.**
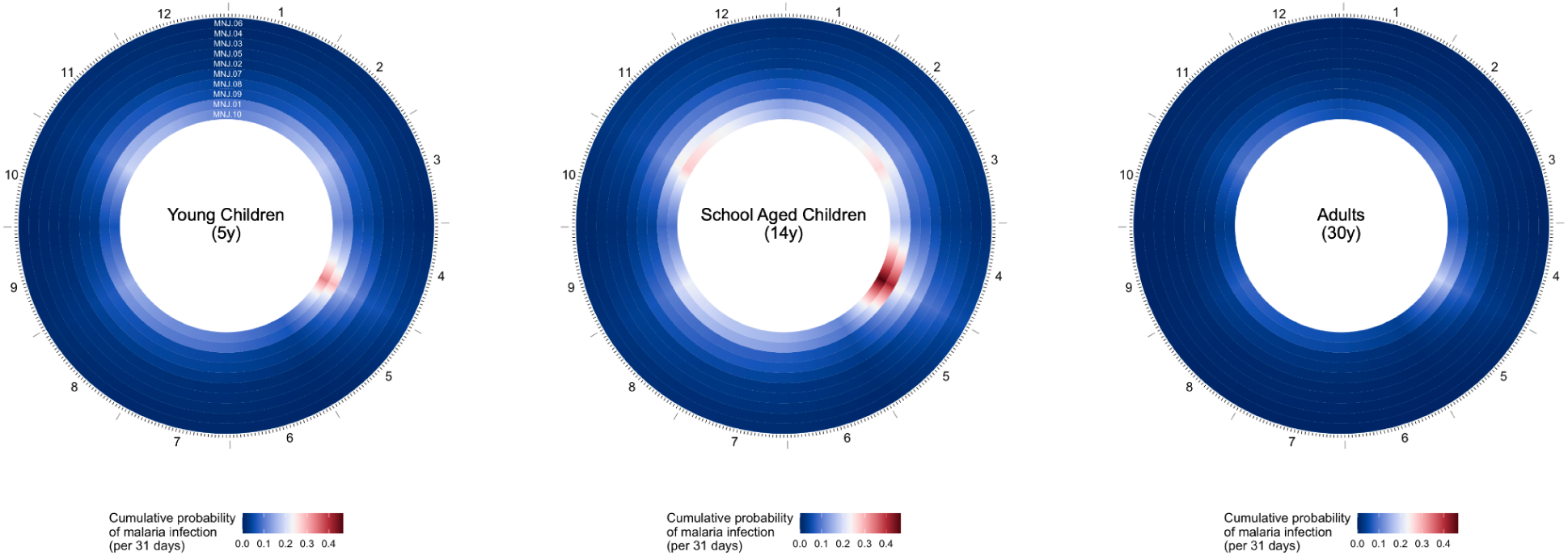
Climate and malaria seasonality in Mananjary district. Rings show sites ordered from lowest (outer) to highest (inner) infection rate for three select age groups: Young children (5 years, at left), school aged children (14 years, center), and adults (30 years, right). Color shows the cumulative probability of malaria infection (i.e., the proportion of susceptible individuals expected to have one or more new RDT detectable infections) for a 31-day interval beginning on that day.

**Figure S11.**
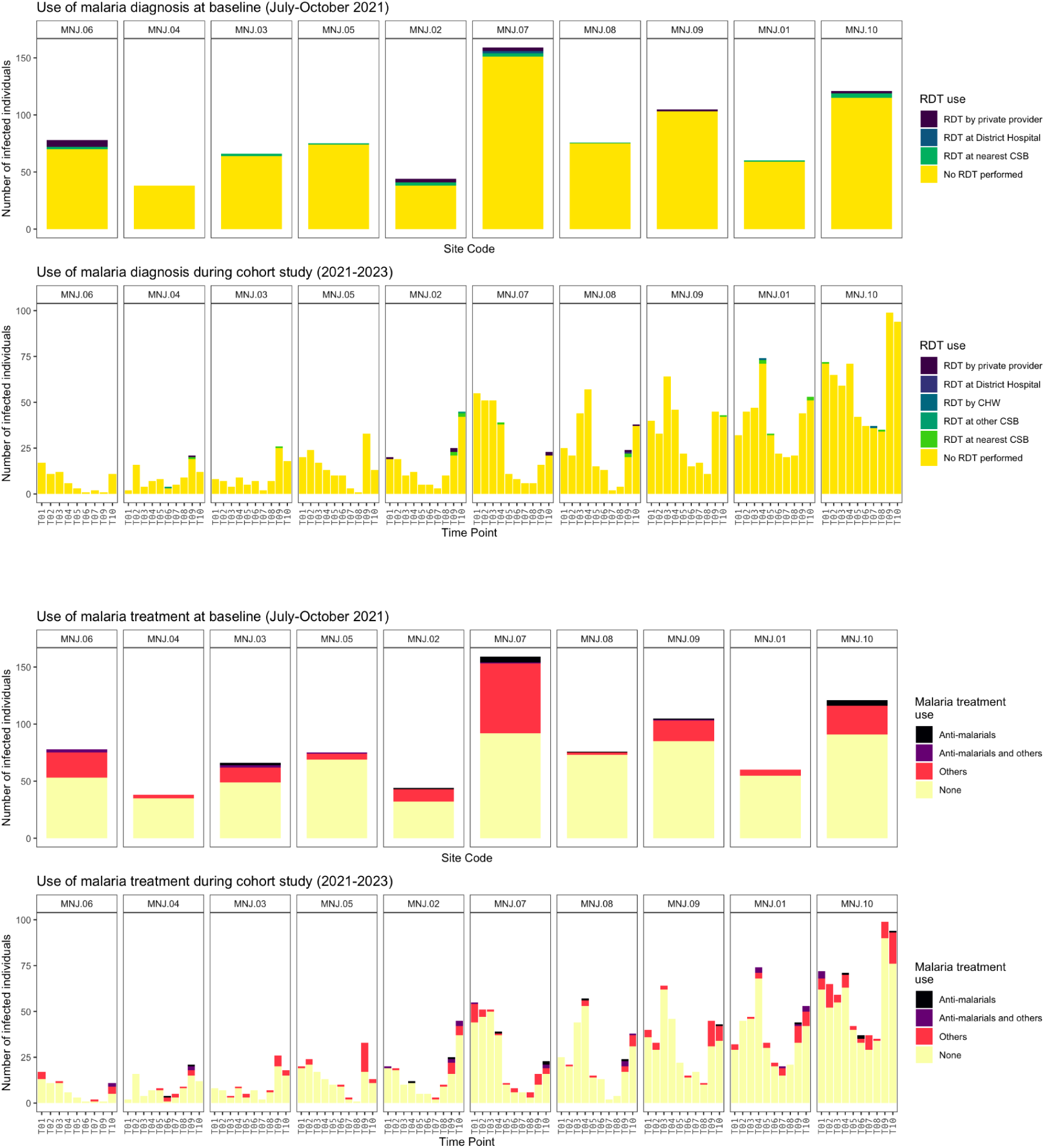
Use of malaria diagnosis and treatment in Mananjary district. Frequency of rapid diagnostic test (RDT) use and anti-malarial use among malaria infected individuals in the Mananjary district cohort sample per recall questionnaire at the time of sampling, by site. Sites ordered by average prevalence from time point T01 to T10. Antimalarials included artemisinin-based combination therapy (ACT) and quinine. CHW: Community Health Worker; CSB: Centres Santé de Bases (rural public health facilities).

**Table S1:**
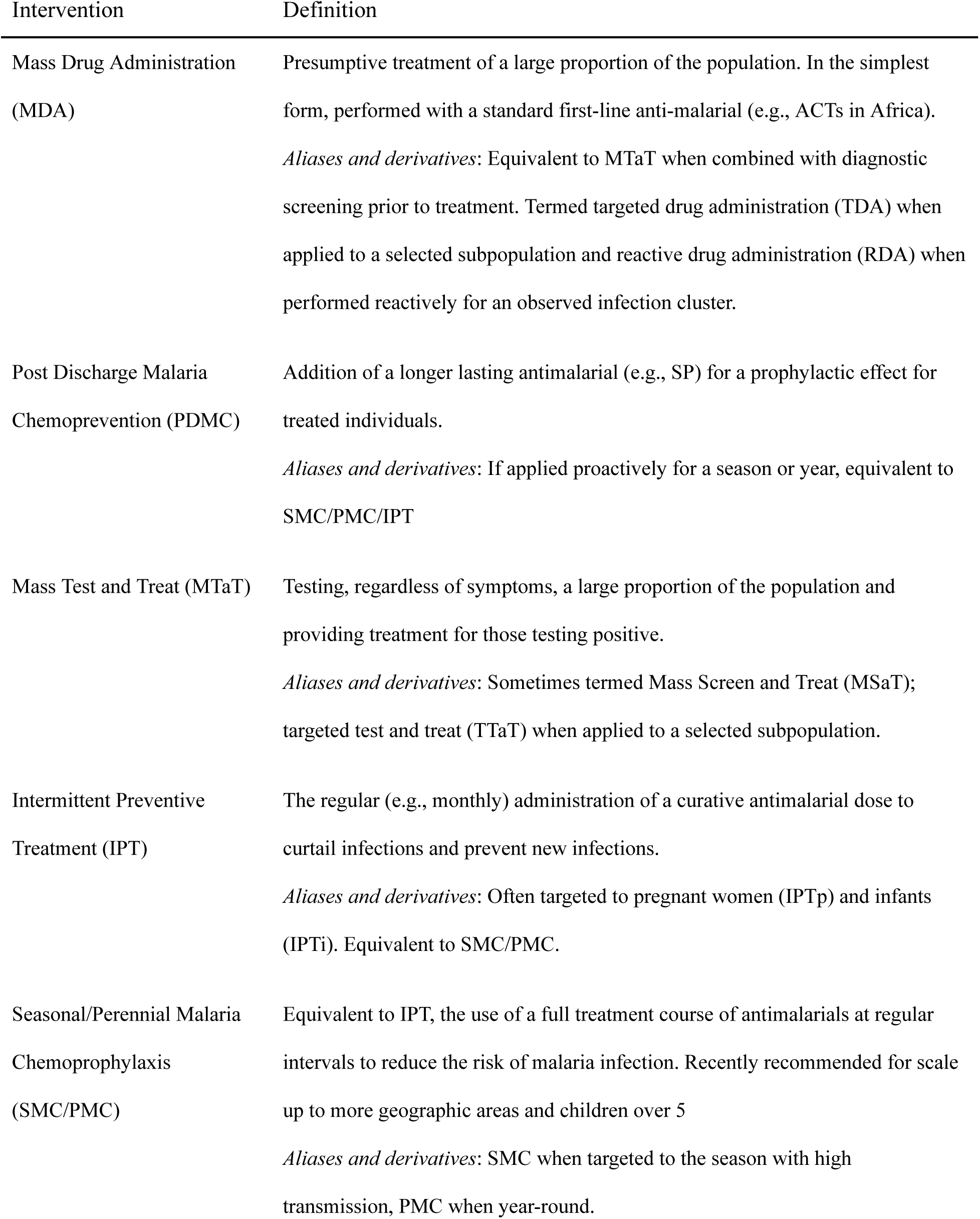
World Health Organization assessed drug-based malaria interventions.

**Table S2.** Parameter definitions used in vaccination modeling.

| Parameter | Definition | Range explored |
| --- | --- | --- |
| $1 - e^{-\lambda t}$ | $p(I)$ = Proportion of uninfected individuals that test positive during interval $t$ | 0.01-0.7 |
| $v$ | Vaccination coverage: the proportion of individuals fully vaccinated at the onset of the period of focus | 0-1 |
| $v_p$ | Partial vaccination coverage: the proportion of individuals partially vaccinated at the onset of the period of focus | 0-1 |
| $s$ | Proportion of infected individuals reporting symptoms | 0.1-0.6 |
| $E_s$ | Efficacy against symptomatic infection for completely vaccinated individuals | 0-1 |
| $E_p$ | Efficacy against symptomatic infection for partially vaccinated individuals | $0-E_s$ |
| $E_i$ | Efficacy against infection | 0 |

